# Border closure and travel restrictions to control the spread of COVID-19: an update to a Cochrane review

**DOI:** 10.1101/2022.01.22.22269686

**Authors:** Ahmed M. Abou-Setta, Otto LT Lam, Viraj Kasireddy, Nicole Askin, Andrea C. Tricco

## Abstract

**Background:** COVID-19 has proven to be more difficult to manage for many reasons including its high infectivity rate. One of the potential ways to limit its spread is by limiting free travel across borders, including via air travel. The objective of this systematic review is to identify, critically-appraise and summarize evidence on border closures and travel restrictions.

**Methods:** This review is based on the Cochrane review: “International travel-related control measures to contain the COVID-19 pandemic” and followed the same methodology. In brief, we searched for clinical and modelling studies in general health and COVID-19-specific bibliographic databases. The primary outcome categories were (i) cases avoided, (ii) cases detected, and (iii) a shift in epidemic development. Secondary outcomes were other infectious disease transmission outcomes, healthcare utilisation, resource requirements and adverse effects if identified in studies assessing at least one primary outcome.

**Results:** We included 43, mostly modelling, studies that met our inclusion criteria. Fourteen new studies were identified in the updated search, as well as updated companions (e.g., peer-reviewed publications that were previously only available as pre-prints). Most studies were of moderate to high quality. The added studies did not change the main conclusions of the Cochrane review nor the quality of the evidence (very low to low certainty). However, it did add to the evidence base for most outcomes.

**Conclusions:** Weak evidence supports the use of border closures to limit the spread of COVID-19 via air travel. Real-world studies are required to support these conclusions.

## Background

In humans, coronaviruses may cause respiratory infections ranging from the common cold to severe disease. The 2003 Severe Acute Respiratory Syndrome (SARS), the 2012 Middle Eastern Respiratory Syndrome (MERS), and the 2019 coronavirus disease (COVID-19) are all notable pandemics caused by coronaviruses.

COVID-19 has proven to be more difficult to manage, compared to previous epidemics, for many reasons including its high infectivity rate. The mean reproductive number (R_0_), which represents the speed of spread or transmissibility, of SARS-CoV-2 (the virus that causes COVID-19) has been estimated to be around 3.28,^1^ which is higher than that for SARS (1.7–1.9) and MERS (<1).^2^

To combat the transmission of SARS-CoV-2, governments and public health organizations/ officials have implemented polices to decrease the disease spread including border closures and travel restrictions. A recent Cochrane review^3^ showed that there was little evidence to demonstrate the effectiveness of such policies.

The objective of this systematic review is to identify, critically-appraise and summarize evidence on border closures and travel restrictions to control the spread of SARS-CoV-2 transmission between countries and regions.

## Methods

As this is an update to a Cochrane review,^3^ we conducted this review according to guidelines enumerated in the Methodological Expectations of Cochrane Intervention Reviews (MECIR), and reported according to the Preferred Reporting Items for Systematic reviews and Meta-Analyses (PRISMA) guidelines.^4^ We included randomized trials, non-randomized trials, observational studies, and modelling studies on the effects of travel related control measures affecting human travel during the COVID 19 pandemic. The interventions for this review were border closure and travel restrictions to control the spread of COVID-19. The non-randomized, observational, and modelling studies could be single arm or with a control group, including but not limited to prospective or retrospective cohort studies, case-controlled studies, cross-sectional studies, interrupted time series, or mathematical modelling studies. Modelling studies could use a theoretical comparison. We excluded case reports/ series, opinion papers, editorials, study protocols and trial registries.

The primary outcome categories were (i) cases avoided, (ii) cases detected, and (iii) a shift in epidemic development. Secondary outcomes were other infectious disease transmission outcomes, healthcare utilisation, resource requirements and adverse effects if identified in studies assessing at least one primary outcome.

### Search strategy for identification of studies

The Cochrane review^3^ search was adapted by excluding terms not related to COVID-19 (e.g., MERS, H1N1, SARS01) and an updated search conducted from Nov 2020 to Jun 2021 without any language restrictions. The search was conducted in general health and COVID-19-specific bibliographic databases [PubMed (NLM), Cochrane Covid (https://covid-19.cochrane.org/), Medrxiv (https://connect.Medrxiv.org/relate/content/181), and BioRXiv. We conducted searches in general purpose databases (e.g., Google), government and public health websites (e.g., WHO), preprint servers (arxiv.org) and news outlets for additional unpublished or grey literature. Each database was searched using an individualized search strategy. Finally, the reference lists of relevant narrative and systematic reviews and included studies were hand-searched for relevant citations. We performed reference management in EndNote™ (version X9, Thomson Reuters, Carlsbad, CA, USA).

### Study selection

We used a two-stage process for study screening and selection using standardized and piloted screening forms. Two reviewers independently screened the titles and abstracts of search results to determine if a citation met the inclusion criteria. Full texts of all included citations were reviewed independently, and in duplicate. All conflicts were resolved through discussion, consensus or by a third researcher, as required.

### Data abstraction and management

One reviewer summarized the findings from included study reports, and a second researcher reviewed the summaries for accuracy and completeness. Discrepancies between the two reviewers were resolved by discussion and consensus. Data management was performed using Microsoft Excel™ 2010 (Excel version 14, Microsoft Corp., Redmond, WA, USA).

### Assessment of methodological quality and potential risk of bias

Evidence from modelling studies was assessed using the tools proposed by Jaime Caro et al., 2014.^5^ Nonrandomized comparative studies were assessed using the Newcastle-Ottawa Scale.

### Data summary

All data is summarized narratively and in tabular format.

## Results

From 4,462 unique citations, we included 43 studies that met our inclusion criteria (Figure 1). In addition to the 31 studies^6–36^ previously identified by the Cochrane review,^3^ in the updated search, we identified 12 new studies^37–48^ that met the inclusion criteria; of which five studies were specific to air travel restrictions/ bans^39, 40, 44, 45, 48^ and seven were general travel restrictions/ bans, including air travel.^37, 38, 41–43, 46, 47^

**Figure 1.**
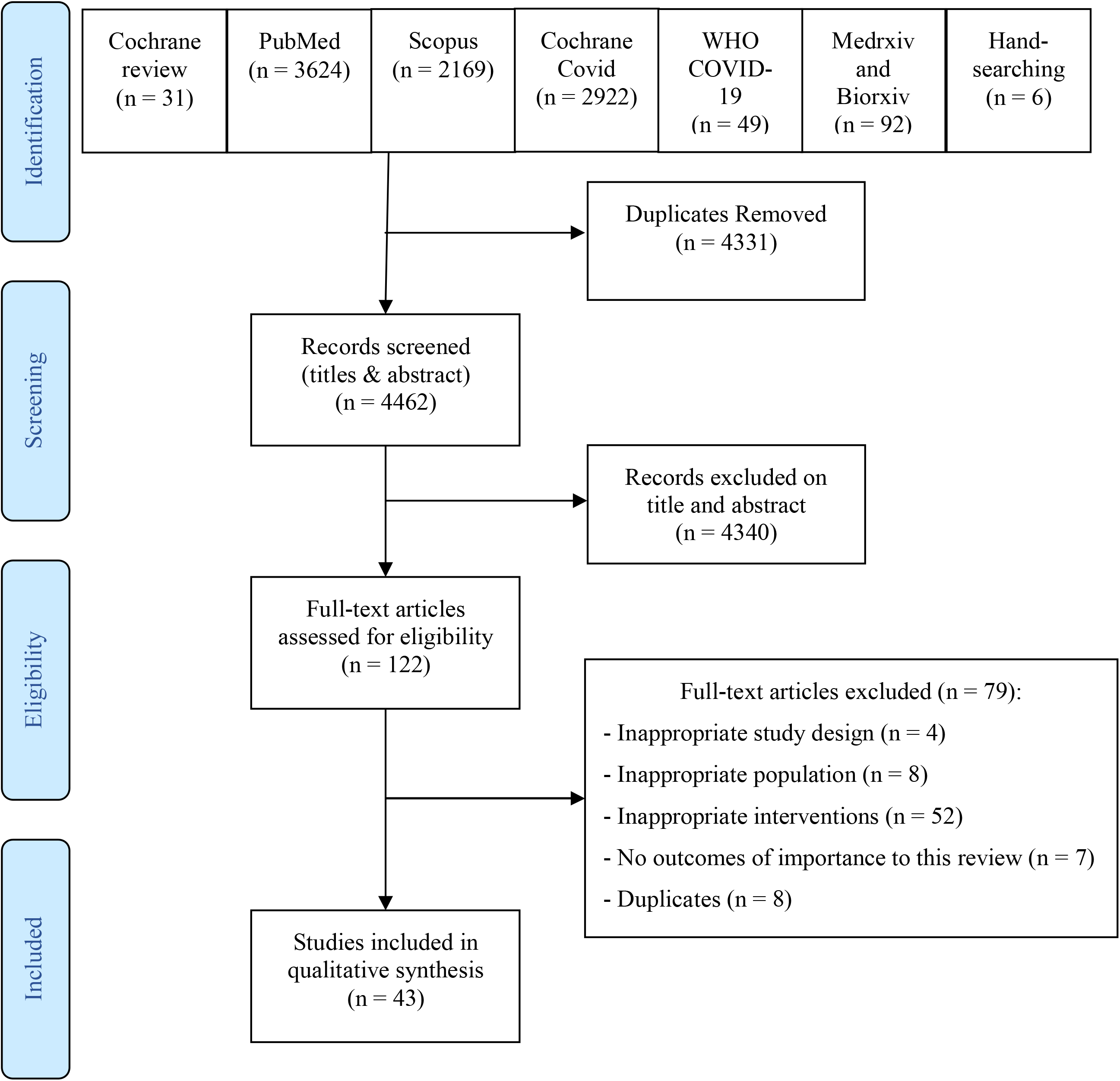
PRISMA Flow Diagram.

### Study designs

We included 35 modelling studies^6–36, 40, 42, 43, 48^ of which four studies^40, 42, 43, 48^ were identified in the updated search. One of the newly included studies was a nonrandomized comparative study.^38^ Neither the Cochrane review^3^ nor the updated search identified any additional observational studies.

### Quality appraisal

None of the included modelling studies fulfilled all the quality appraisal items; all had moderate to major concerns. Fifty-seven percent of the studies (n = 24) fulfilled (i.e., no to minor concerns) at least 50% of the items. The most concerns were regarding internal validation: “Has the internal validation process been described?”, “Has the model been shown to be internally valid?” and “Was technical documentation, in sufficient detail to allow (potentially) for replication, made available openly or under agreements that protect intellectual property?”. The quality assessments for the included studies are summarized in Table 3 and 4, with individual study assessment for the studies identified in the updated search presented in Appendix 1.

**Table 1.** Study characteristics of studies identified in the updated search.

|  | **Gankin 2021** | **Gwee 2021** | **Han 2021** | **Hossain 2020** | **Ip 2021** |
| --- | --- | --- | --- | --- | --- |
| **Study design:** | Modelling study | Comparative study | Modelling study | Modelling study | Mathematical modeling study |
| **Description:** | Epidemiological study analyzing the characteristics of the first months of the epidemic in Ukraine using agent-based modelling and phylodynamics. | Comparative analysis was conducted on Singapore, Taiwan, Hong Kong and South Korea's COVID-19 response. Data on COVID-19 cases, travel-related and community interventions, socio-economic profile were consolidated. Trends on imported and local cases were analyzed using computations of moving averages, rate of change, particularly in response to distinct waves of travel-related interventions due to the outbreak in China, South Korea, Iran & Italy, and Europe. | This modeling study included flight routes between 48 origins, including Hong Kong, Macau, and Taiwan in China and 45 foreign countries, and 44 destination cities in China with international airports. The daily number of international flights for each origin-destination pair were aggregated. A destination- and time-specific abroad infection index was constructed, and a counterfactual analysis was performed. | SIR meta-population model with a mobility matrix (contact mixing at the population level). The SIR model was embedded with analytical expressions that could stratify the imported cases and the secondary cases produced by the imported cases along with other infected individuals with border control and quarantine measures. | This modeling study assessed policies introduced by the Australian government using a generalised space-time autoregressive model which incorporated multiple exogenous variables and delay effects. |
| **Travel-related control measure(s):** | Border closure | Border closure, quarantine | Mandatory testing at customs, centralized quarantine, and a ban on the entry of foreigners and the "five one" international flight restrictions | Border control and quarantine measures | International border restrictions |
| **Country implementing measure(s):** | Ukraine | Singapore, Taiwan, Hong Kong, South Korea | China | Wuhan, China | Australia |
| **Outcome(s):** | Time to outbreak, Number or proportion of cases in the community | Proportion of imported cases, number or proportion of cases in the community | Abroad infection index | Outbreak arrival time | Number of cases |
| **Follow-up:** | Mar 12 to Aug 1, 2020 | Mar to Apr 5, 2020 | Mar 18 to Apr 2, 2020 | 14 days | A "few days" after policy implementation |
| **Funding:** | Non-industry | Non-industry | Non-industry | Non-industry | Non-industry funding |
| **COI** | No COIs reported | No COIs reported | No COIs reported | No COIs reported | No COIs reported |

|  | **Jablonska 2021** | **Kwak 2021** | **Sun 2021** | **Yang 2021** | **Yang 2021** |
| --- | --- | --- | --- | --- | --- |
| **Study design:** | Modelling study | Modelling study | Mathematical modeling study | Modeling study | Modeling study |
| **Description:** | Univariate and multivariate generalised linear models using different selection algorithms (forward, backward, stepwise, and genetic algorithm) | Deep reinforcement learning along with dueling Double Deep Q-Network algorithm were used | Network based SEIR model, inspired by the Global Epidemic and Mobility Model, was used to analyze flight data for 2020-2021 to determine whether existing flight bans were effective against newly emerged variants of concern. | A Bayesian framework was used to model disease progress of COVID-19 and the effectiveness of travel measures using data on passengers arriving in Hong Kong and laboratory-confirmed imported cases. | Data from 165 countries was analyzed to determine the effectiveness of international travel controls in delaying epidemic peaks. |
| **Travel-related control measure(s):** | Travel restrictions reducing or stopping cross-border travel | International travel restrictions by difference in policy levels between the government and agent | Flight bans | Travel bans for non-Hong Kong residents from overseas. Testing and quarantine for those permitted to travel. | Closing borders, screening of inbound travelers, quarantine |
| **Country implementing measure(s):** | 34 countries in Europe | 216 countries | China, India, USA, Pakistan, Indonesia, Brazil, Mexico, France, UK, Italy, Germany, Japan, Russia, Nigeria, Bangladesh, Ethiopia, Philippines, Egypt, Vietnam, Congo, Turkey, Thailand, Iran, South Africa, Tanzania | Hong Kong | 165 countries |
| **Outcome(s):** | Time to outbreak, number of deaths at peak | Proportion of cases detected | Delay in arrival time of COVID-19 variant | Imported cases, percentage of infectious travellers mixing with the community, risk of releasing infectious travelers | Time to outbreak |
| **Follow-up:** | Not reported | Jan 2020 to Nov 2020 | 2020-2021 | April 1 to July 31, 2020 | Time from January 1, 2020, to the first epidemic peak (which was identified from the modal daily case counts within any 53-day sliding window) |
| **Funding:** | Not funded | Not funded | Non-industry funding | Non-industry funding | Non-industry funding |
| **COI** | No COIs reported | No COIs reported | No COIs reported | Industry consultant | Industry consultant |

|  | Yu 2021 | Zhu 2021 |
| --- | --- | --- |
| **Study design:** | Modeling study | Modelling study |
| **Description:** | This study reconstructed COVID-19 transmission from historical data and simulated the effects of three different border-reopening policies. | SUIHR model with the following features: 1) explicit modeling of the importation of infected individuals from one entity to another, to facilitate the assessment of the effect of border control policies; 2) model focused on the undetected spreading of virus from mainly presymptomatic or asymptomatic people (The Infectious state in the SIR models were deliberately split into unidentified Infectious and Identified Infectious states, to account for the characteristics of a reopening phase compared to the initial outbreak control phase); 3) contact tracing was built into the model (during a reopening phase, "trace & isolate" was preferred over lockdowns as it is less disturbing to daily life and hence more sustainable); and 4) constraints such as new case targets and medical resources were incorporated into the model framework as part of the policy optimization process. Solutions to the model were obtained via linear programming. |
| **Travel-related control measure(s):** | Daily quota for inbound travelers, contact tracing | Border control policies, in combination with internal measures (model with built-in imported risk and (1-tier) contact tracing) |
| **Country implementing measure(s):** | Hong Kong | G1 to G3 countries |
| **Outcome(s):** | Cumulative cases during first 60 days | Delay of outbreak, weekly new identified cases |
| **Follow-up:** | 60 days | Not reported |
| **Funding:** | Non-industry funding | Not reported |
| **COI** | No COIs reported | No COIs reported |

COI: Conflicts of interest; SIR: Susceptible, Infected, and Recovered

**Table 2.**
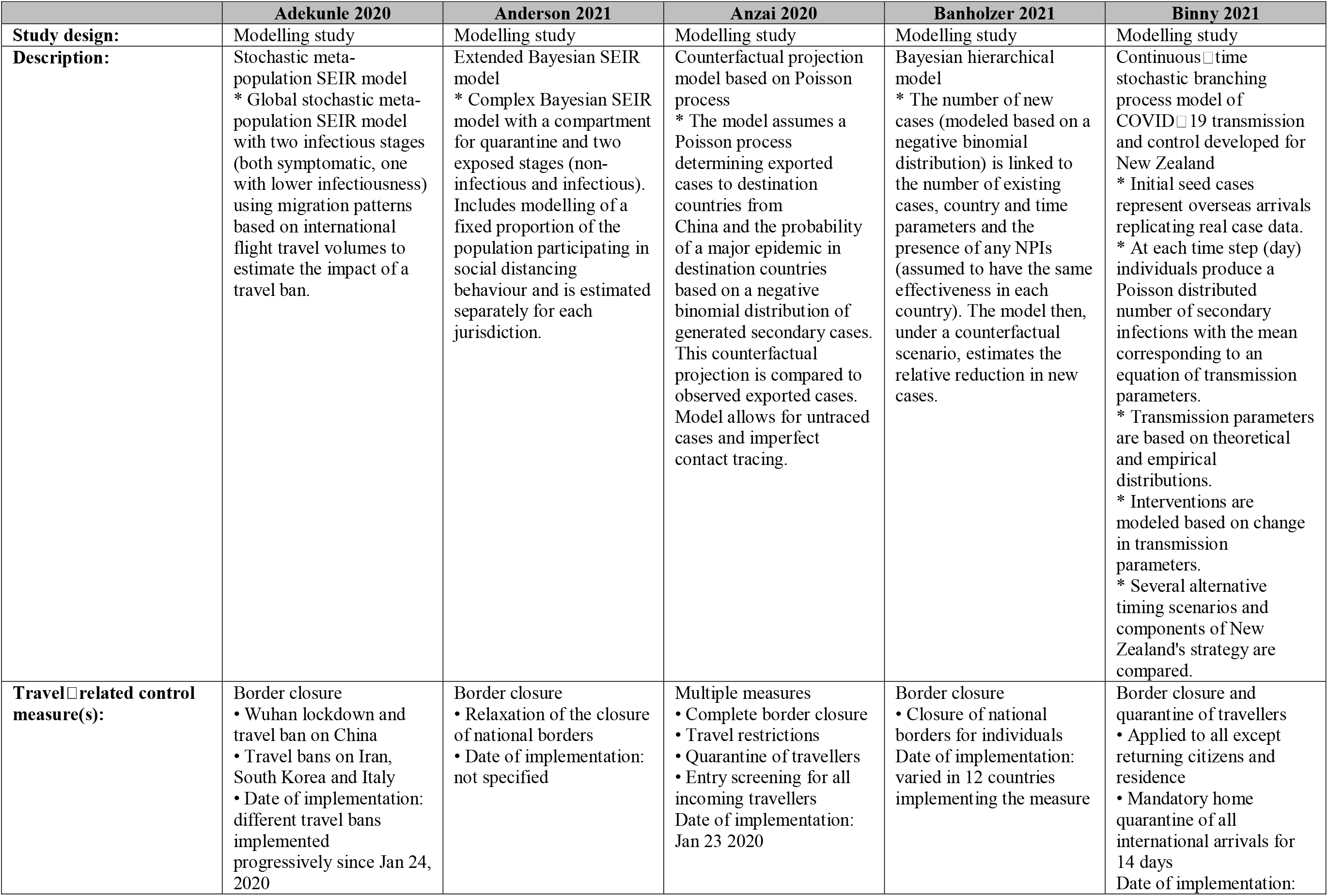
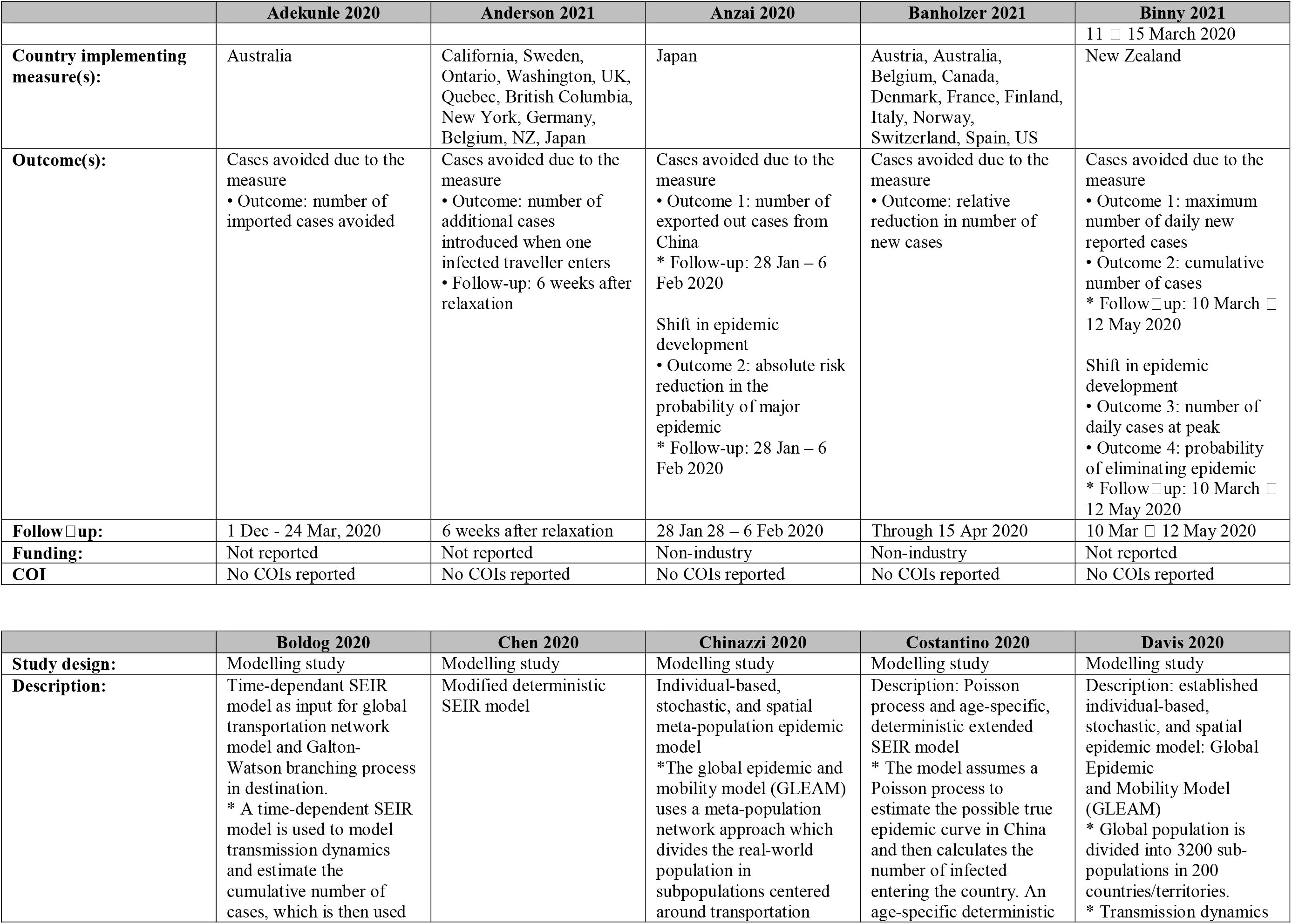
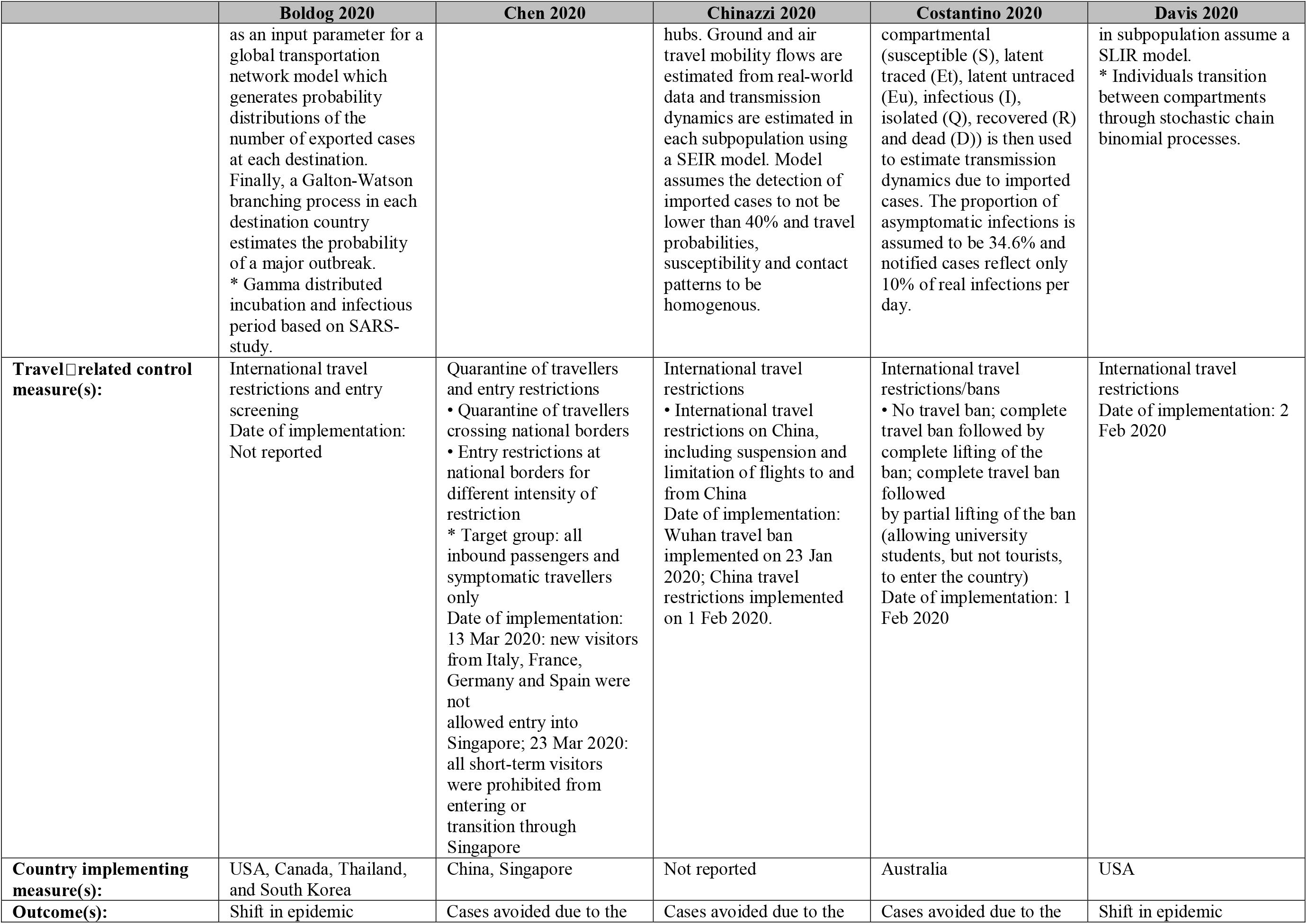
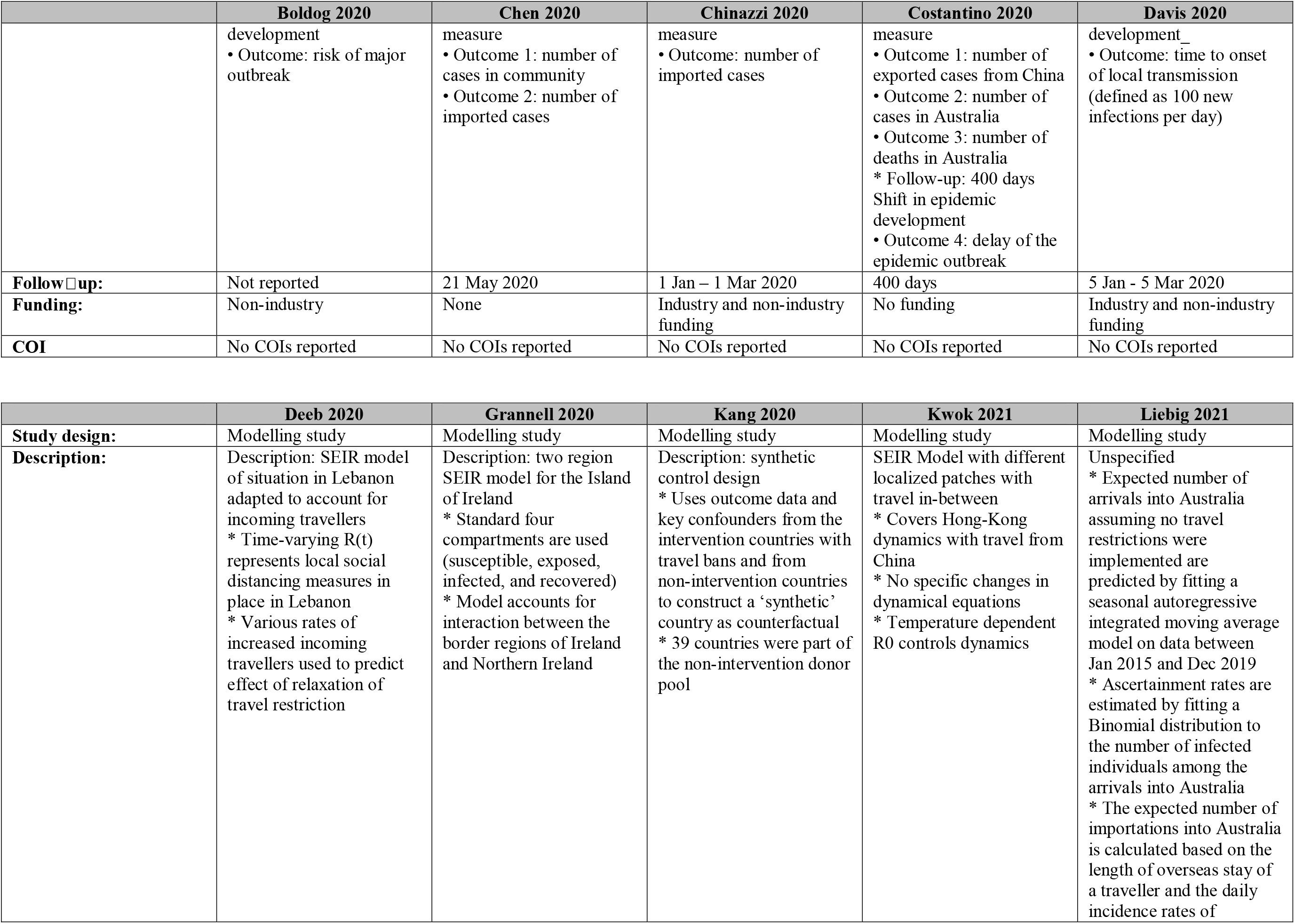
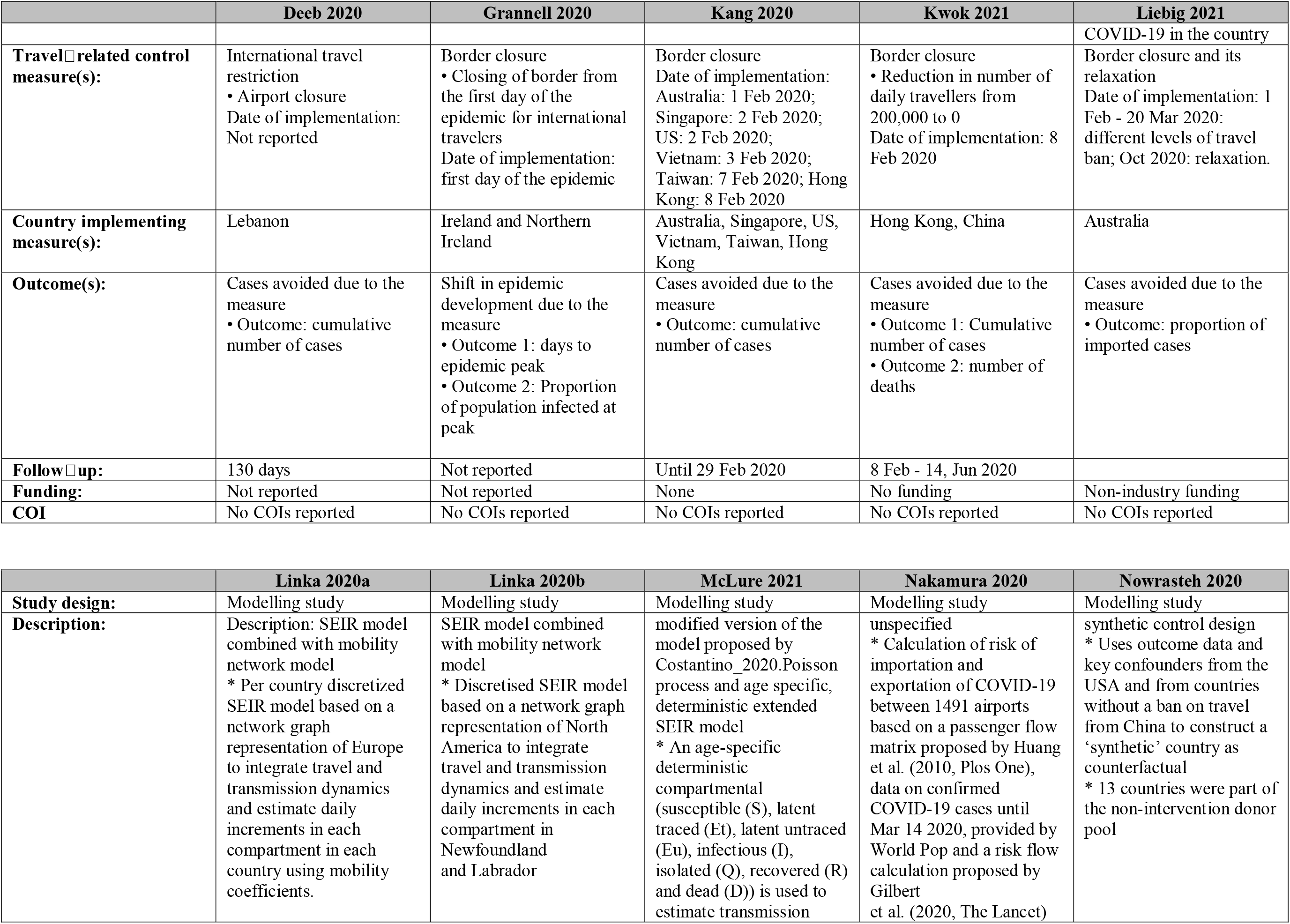
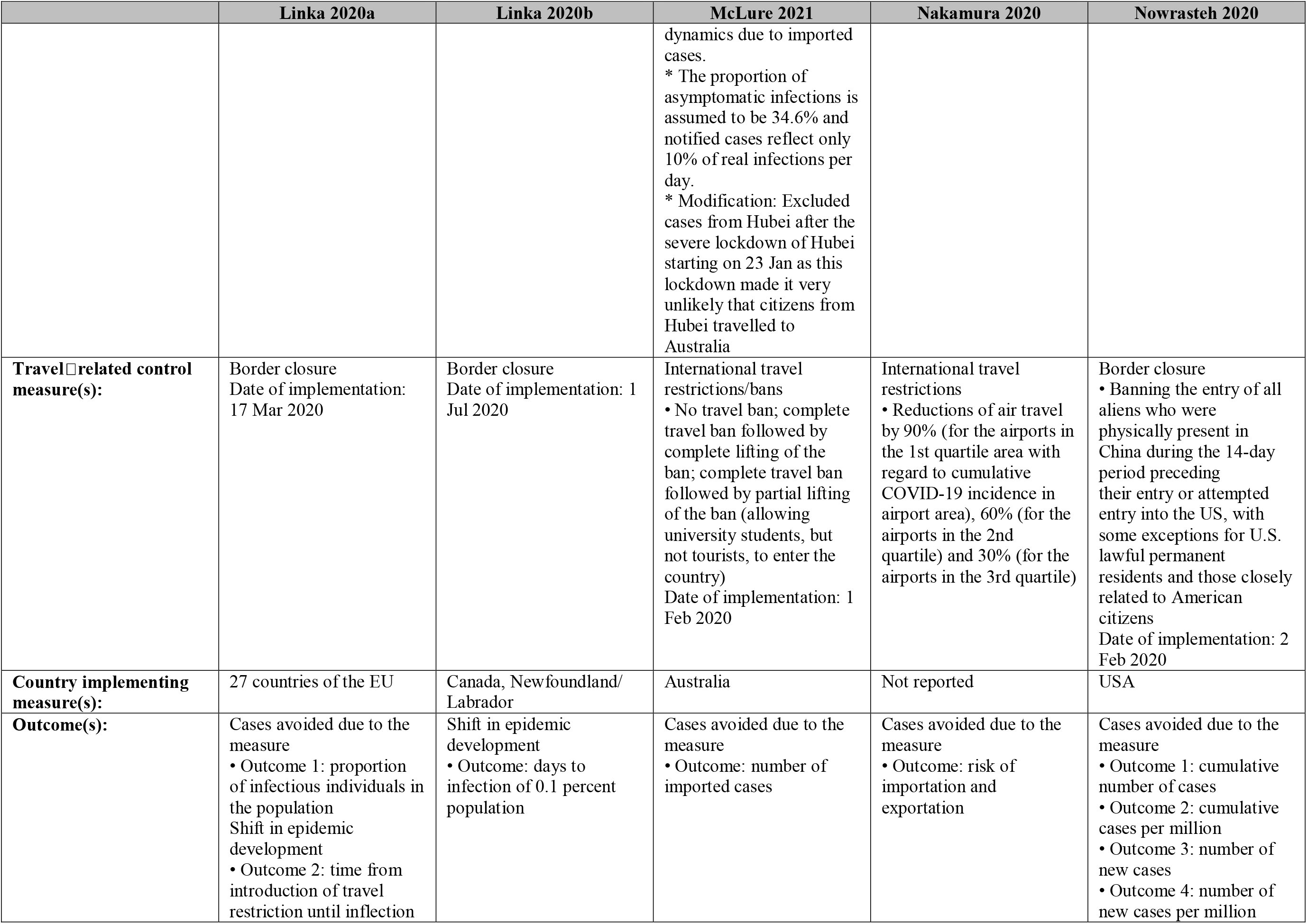
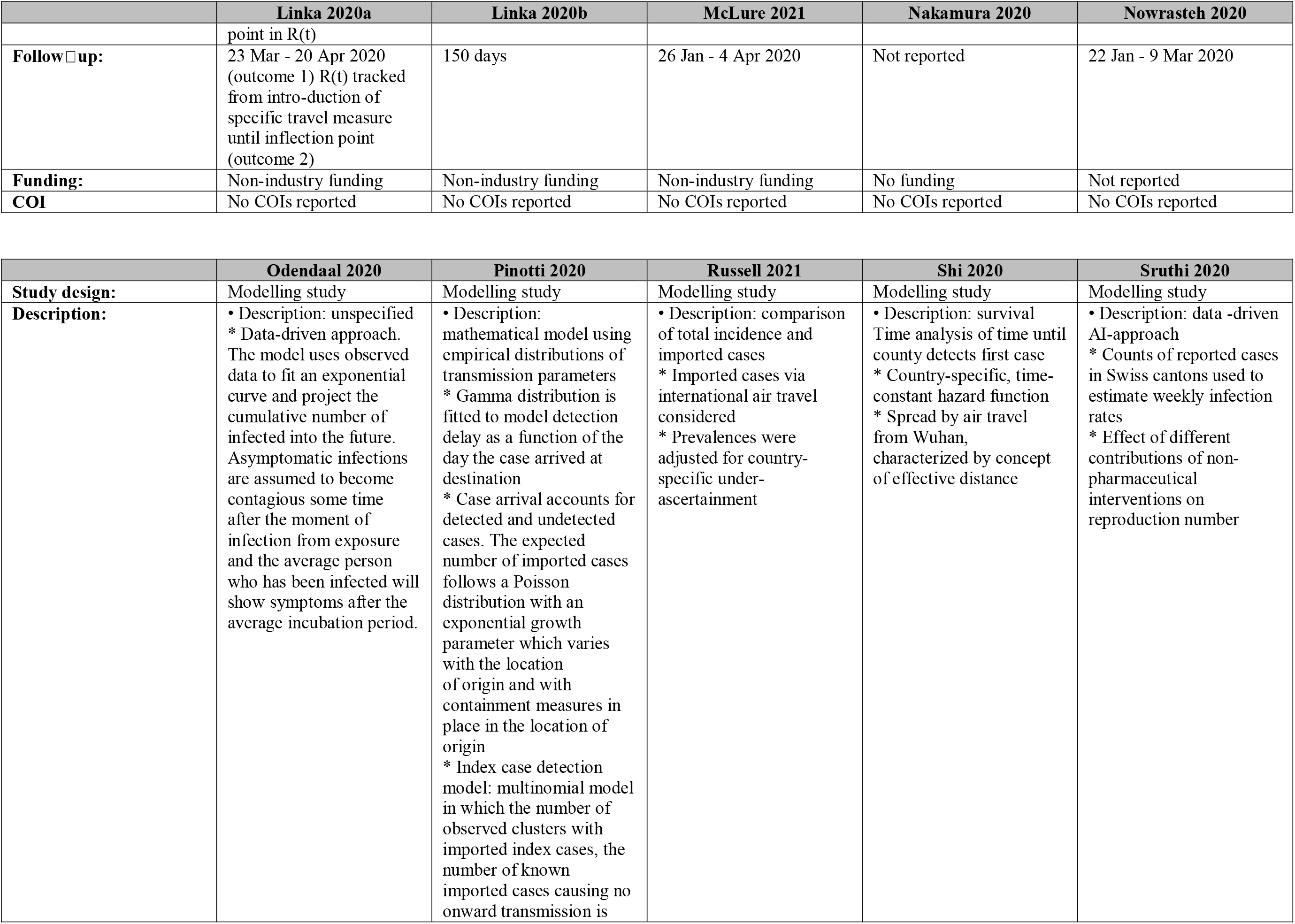
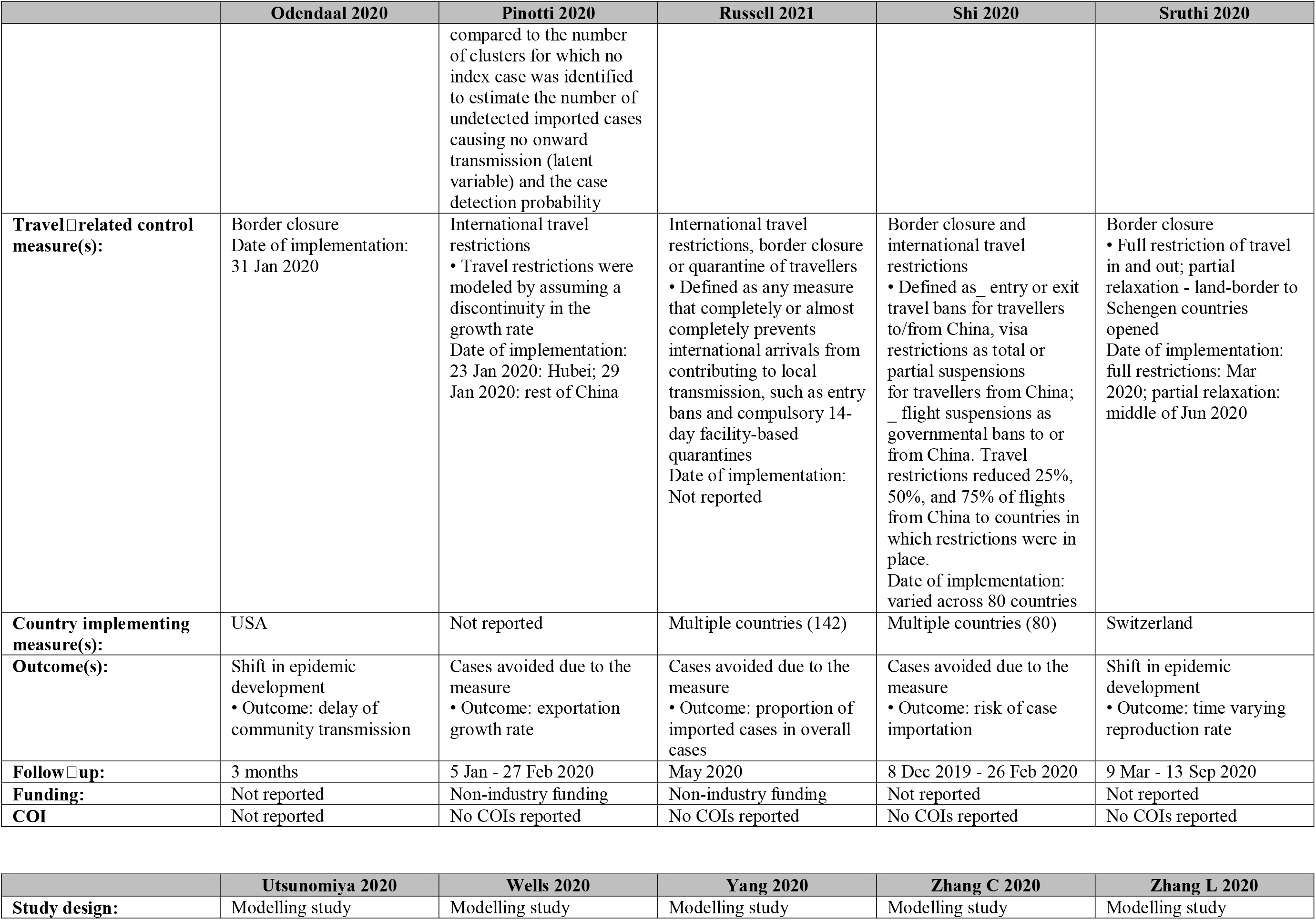
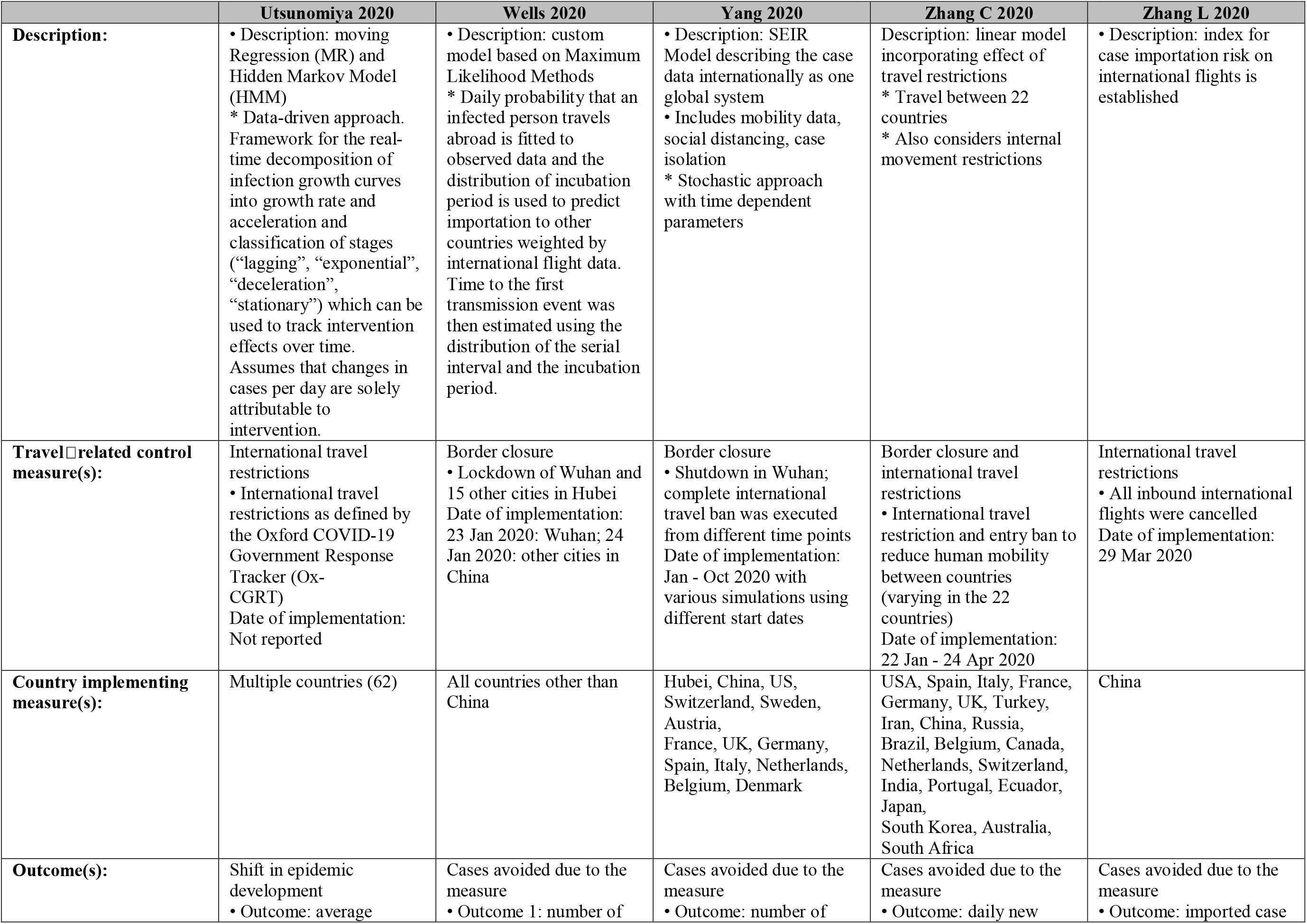
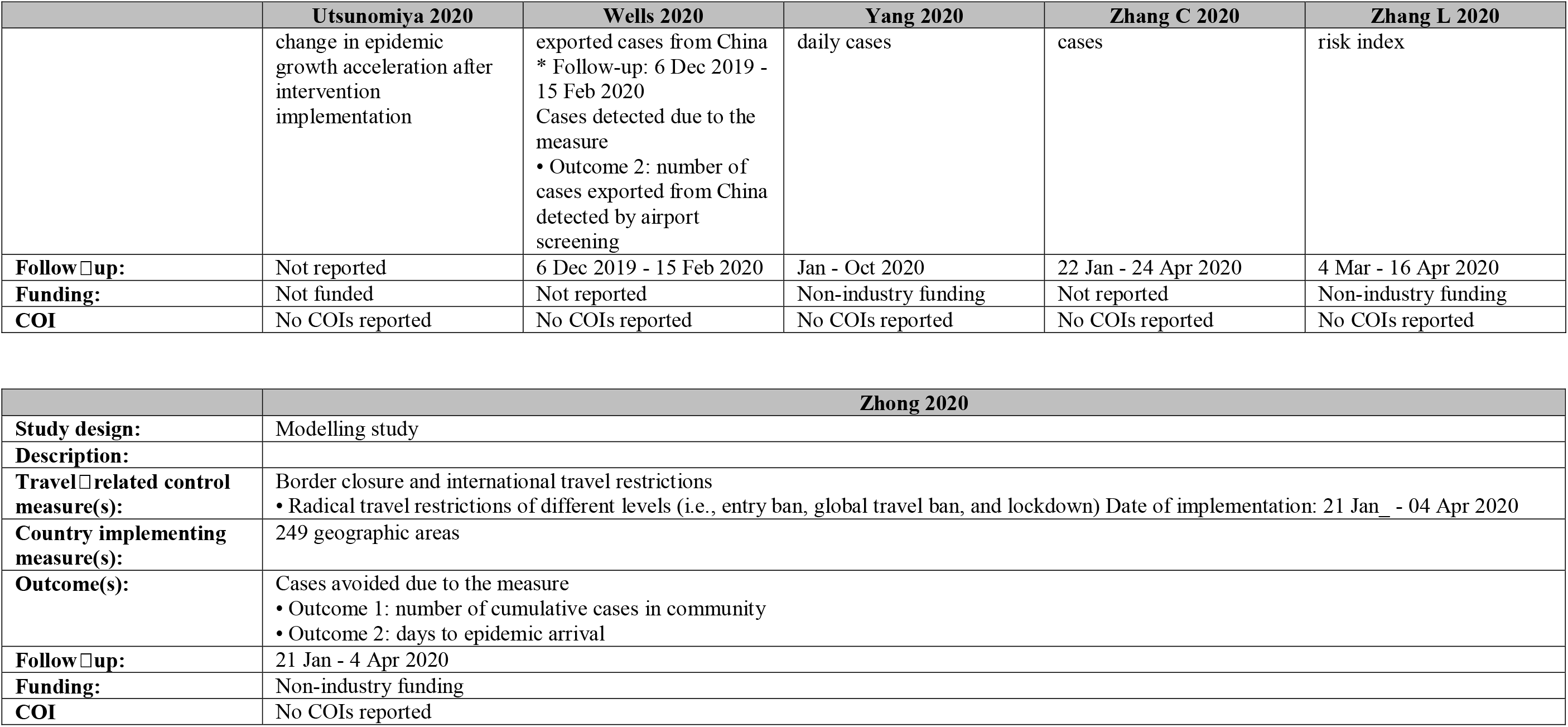
Study characteristics of studies identified in the original Cochrane review.

**Table 3.**
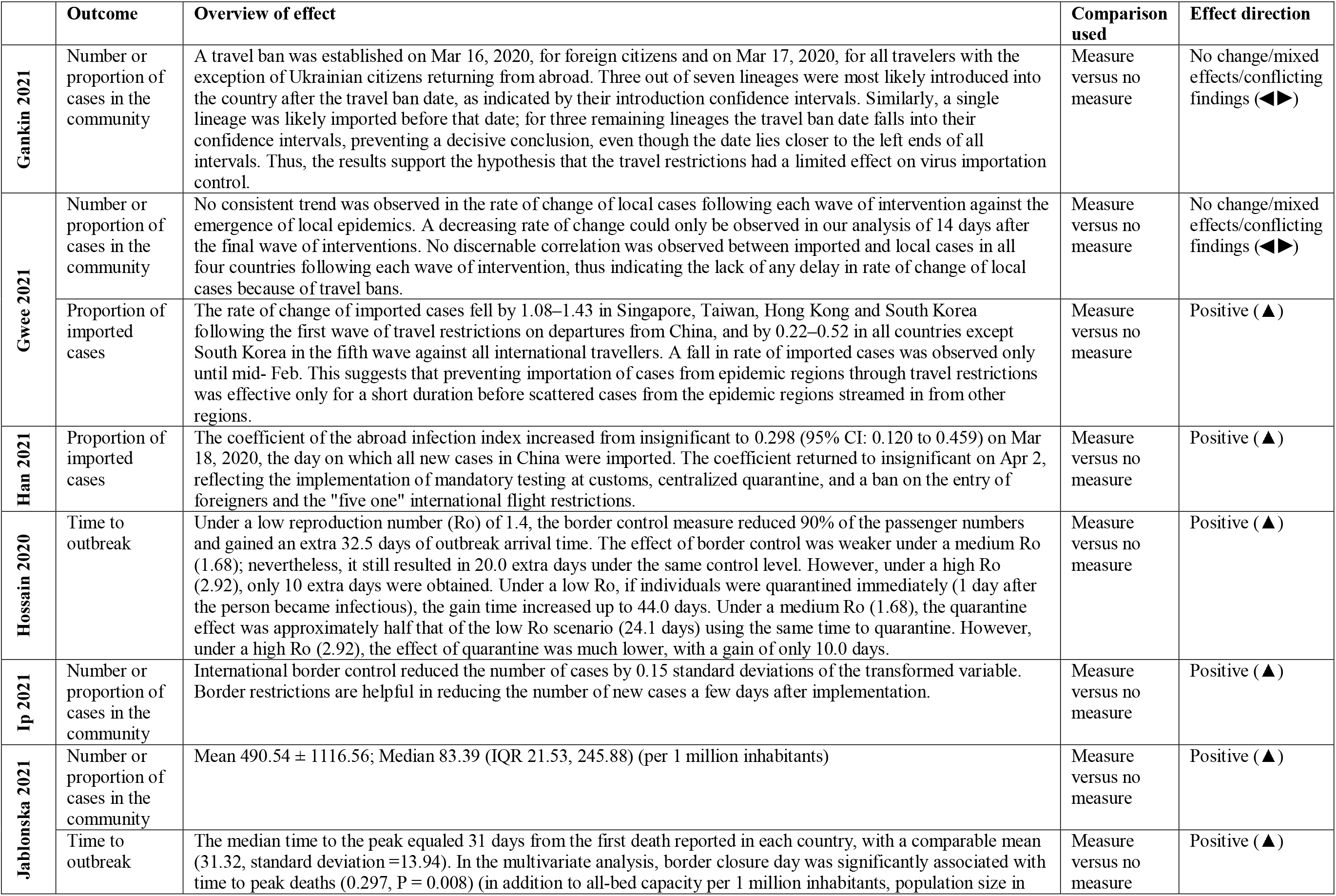
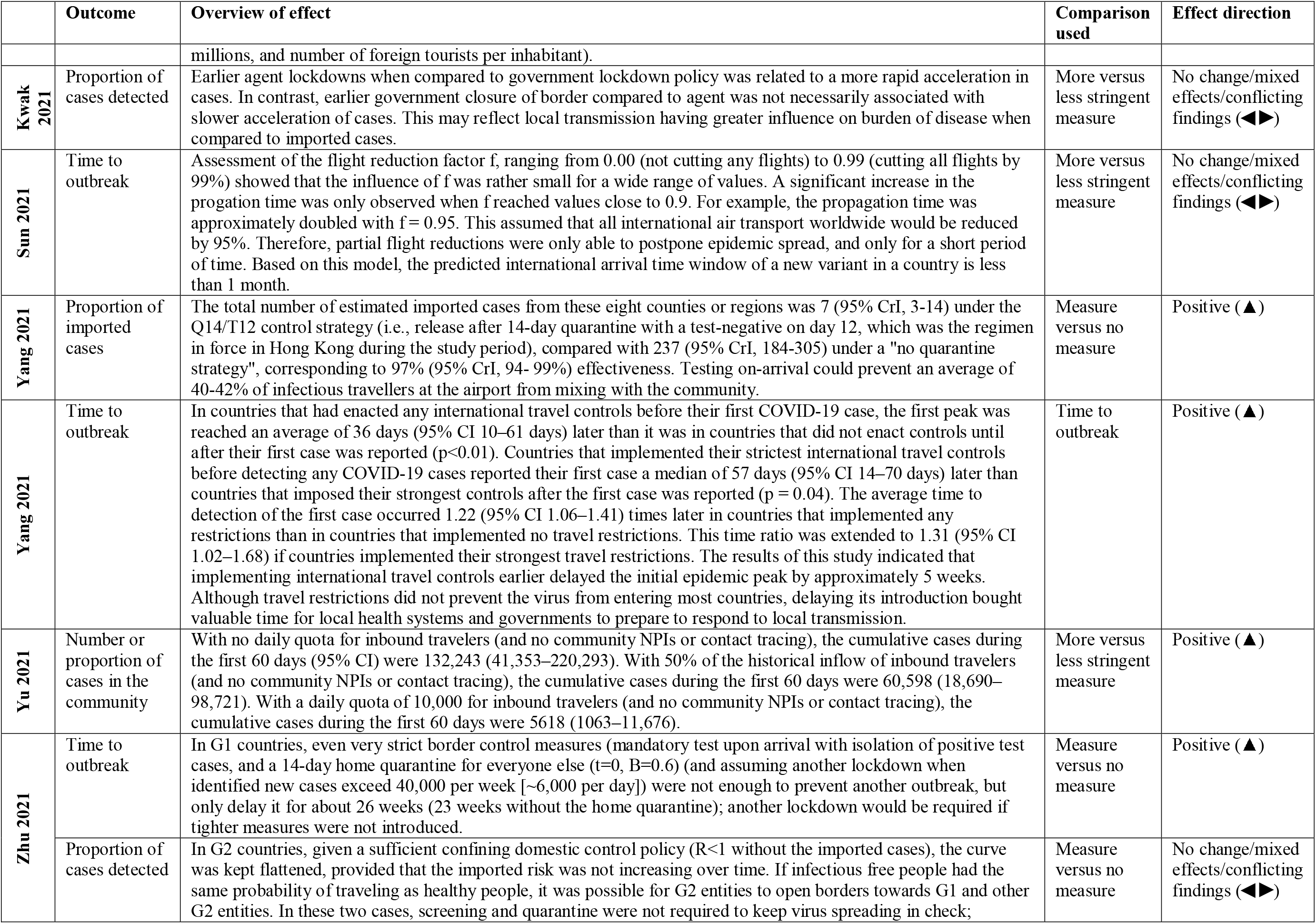
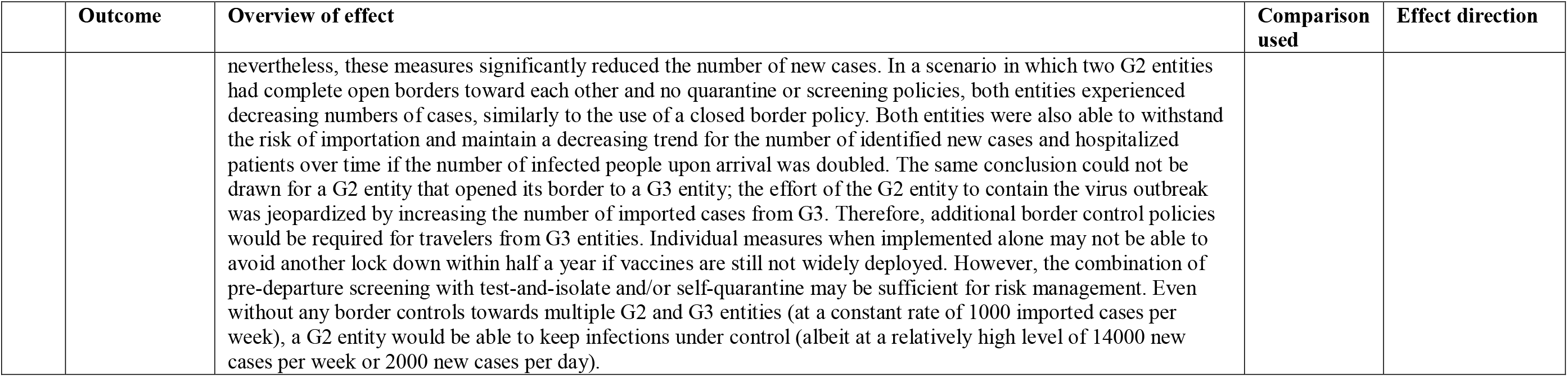
Outcomes of studies identified in the updated search.

|  | Outcome | Overview of effect | Comparison used | Effect direction |
| --- | --- | --- | --- | --- |
| Gankin 2021 | Number or proportion of cases in the community | A travel ban was established on Mar 16, 2020, for foreign citizens and on Mar 17, 2020, for all travelers with the exception of Ukrainian citizens returning from abroad. Three out of seven lineages were most likely introduced into the country after the travel ban date, as indicated by their introduction confidence intervals. Similarly, a single lineage was likely imported before that date; for three remaining lineages the travel ban date falls into their confidence intervals, preventing a decisive conclusion, even though the date lies closer to the left ends of all intervals. Thus, the results support the hypothesis that the travel restrictions had a limited effect on virus importation control. | Measure versus no measure | No change/mixed effects/conflicting findings (◄►) |
| Gwee 2021 | Number or proportion of cases in the community | No consistent trend was observed in the rate of change of local cases following each wave of intervention against the emergence of local epidemics. A decreasing rate of change could only be observed in our analysis of 14 days after the final wave of interventions. No discernable correlation was observed between imported and local cases in all four countries following each wave of intervention, thus indicating the lack of any delay in rate of change of local cases because of travel bans. | Measure versus no measure | No change/mixed effects/conflicting findings (◄►) |
|  | Proportion of imported cases | The rate of change of imported cases fell by 1.08–1.43 in Singapore, Taiwan, Hong Kong and South Korea following the first wave of travel restrictions on departures from China, and by 0.22–0.52 in all countries except South Korea in the fifth wave against all international travellers. A fall in rate of imported cases was observed only until mid- Feb. This suggests that preventing importation of cases from epidemic regions through travel restrictions was effective only for a short duration before scattered cases from the epidemic regions streamed in from other regions. | Measure versus no measure | Positive (▲) |
| Han 2021 | Proportion of imported cases | The coefficient of the abroad infection index increased from insignificant to 0.298 (95% CI: 0.120 to 0.459) on Mar 18, 2020, the day on which all new cases in China were imported. The coefficient returned to insignificant on Apr 2, reflecting the implementation of mandatory testing at customs, centralized quarantine, and a ban on the entry of foreigners and the "five one" international flight restrictions. | Measure versus no measure | Positive (▲) |
| Hossain 2020 | Time to outbreak | Under a low reproduction number (Ro) of 1.4, the border control measure reduced 90% of the passenger numbers and gained an extra 32.5 days of outbreak arrival time. The effect of border control was weaker under a medium Ro (1.68); nevertheless, it still resulted in 20.0 extra days under the same control level. However, under a high Ro (2.92), only 10 extra days were obtained. Under a low Ro, if individuals were quarantined immediately (1 day after the person became infectious), the gain time increased up to 44.0 days. Under a medium Ro (1.68), the quarantine effect was approximately half that of the low Ro scenario (24.1 days) using the same time to quarantine. However, under a high Ro (2.92), the effect of quarantine was much lower, with a gain of only 10.0 days. | Measure versus no measure | Positive (▲) |
| Ip 2021 | Number or proportion of cases in the community | International border control reduced the number of cases by 0.15 standard deviations of the transformed variable. Border restrictions are helpful in reducing the number of new cases a few days after implementation. | Measure versus no measure | Positive (▲) |
| Jablonska 2021 | Number or proportion of cases in the community | Mean 490.54 ± 1116.56; Median 83.39 (IQR 21.53, 245.88) (per 1 million inhabitants) | Measure versus no measure | Positive (▲) |
|  | Time to outbreak | The median time to the peak equaled 31 days from the first death reported in each country, with a comparable mean (31.32, standard deviation =13.94). In the multivariate analysis, border closure day was significantly associated with time to peak deaths (0.297, P = 0.008) (in addition to all-bed capacity per 1 million inhabitants, population size in millions, and number of foreign tourists per inhabitant). | Measure versus no measure | Positive (▲) |
| Kwak 2021 | Proportion of cases detected | Earlier agent lockdowns when compared to government lockdown policy was related to a more rapid acceleration in cases. In contrast, earlier government closure of border compared to agent was not necessarily associated with slower acceleration of cases. This may reflect local transmission having greater influence on burden of disease when compared to imported cases. | More versus less stringent measure | No change/mixed effects/conflicting findings (◄►) |
| Sun 2021 | Time to outbreak | Assessment of the flight reduction factor $f$, ranging from 0.00 (not cutting any flights) to 0.99 (cutting all flights by 99%) showed that the influence of $f$ was rather small for a wide range of values. A significant increase in the propagation time was only observed when $f$ reached values close to 0.9. For example, the propagation time was approximately doubled with $f = 0.95$. This assumed that all international air transport worldwide would be reduced by 95%. Therefore, partial flight reductions were only able to postpone epidemic spread, and only for a short period of time. Based on this model, the predicted international arrival time window of a new variant in a country is less than 1 month. | More versus less stringent measure | No change/mixed effects/conflicting findings (◄►) |
| Yang 2021 | Proportion of imported cases | The total number of estimated imported cases from these eight counties or regions was 7 (95% CrI, 3-14) under the Q14/T12 control strategy (i.e., release after 14-day quarantine with a test-negative on day 12, which was the regimen in force in Hong Kong during the study period), compared with 237 (95% CrI, 184-305) under a "no quarantine strategy", corresponding to 97% (95% CrI, 94- 99%) effectiveness. Testing on-arrival could prevent an average of 40-42% of infectious travellers at the airport from mixing with the community. | Measure versus no measure | Positive (▲) |
| Yang 2021 | Time to outbreak | In countries that had enacted any international travel controls before their first COVID-19 case, the first peak was reached an average of 36 days (95% CI 10–61 days) later than it was in countries that did not enact controls until after their first case was reported ($p < 0.01$). Countries that implemented their strictest international travel controls before detecting any COVID-19 cases reported their first case a median of 57 days (95% CI 14–70 days) later than countries that imposed their strongest controls after the first case was reported ($p = 0.04$). The average time to detection of the first case occurred 1.22 (95% CI 1.06–1.41) times later in countries that implemented any restrictions than in countries that implemented no travel restrictions. This time ratio was extended to 1.31 (95% CI 1.02–1.68) if countries implemented their strongest travel restrictions. The results of this study indicated that implementing international travel controls earlier delayed the initial epidemic peak by approximately 5 weeks. Although travel restrictions did not prevent the virus from entering most countries, delaying its introduction bought valuable time for local health systems and governments to prepare to respond to local transmission. | Time to outbreak | Positive (▲) |
| Yu 2021 | Number or proportion of cases in the community | With no daily quota for inbound travelers (and no community NPIs or contact tracing), the cumulative cases during the first 60 days (95% CI) were 132,243 (41,353–220,293). With 50% of the historical inflow of inbound travelers (and no community NPIs or contact tracing), the cumulative cases during the first 60 days were 60,598 (18,690–98,721). With a daily quota of 10,000 for inbound travelers (and no community NPIs or contact tracing), the cumulative cases during the first 60 days were 5618 (1063–11,676). | More versus less stringent measure | Positive (▲) |
| Zhu 2021 | Time to outbreak | In G1 countries, even very strict border control measures (mandatory test upon arrival with isolation of positive test cases, and a 14-day home quarantine for everyone else ($t=0$, $B=0.6$) (and assuming another lockdown when identified new cases exceed 40,000 per week [$\sim 6,000$ per day]) were not enough to prevent another outbreak, but only delay it for about 26 weeks (23 weeks without the home quarantine); another lockdown would be required if tighter measures were not introduced. | Measure versus no measure | Positive (▲) |
| | Proportion of cases detected | In G2 countries, given a sufficient confining domestic control policy ($R < 1$ without the imported cases), the curve was kept flattened, provided that the imported risk was not increasing over time. If infectious free people had the same probability of traveling as healthy people, it was possible for G2 entities to open borders towards G1 and other G2 entities. In these two cases, screening and quarantine were not required to keep virus spreading in check; | Measure versus no measure | No change/mixed effects/conflicting findings (◄►) |
|  |  | nevertheless, these measures significantly reduced the number of new cases. In a scenario in which two G2 entities had complete open borders toward each other and no quarantine or screening policies, both entities experienced decreasing numbers of cases, similarly to the use of a closed border policy. Both entities were also able to withstand the risk of importation and maintain a decreasing trend for the number of identified new cases and hospitalized patients over time if the number of infected people upon arrival was doubled. The same conclusion could not be drawn for a G2 entity that opened its border to a G3 entity; the effort of the G2 entity to contain the virus outbreak was jeopardized by increasing the number of imported cases from G3. Therefore, additional border control policies would be required for travelers from G3 entities. Individual measures when implemented alone may not be able to avoid another lock down within half a year if vaccines are still not widely deployed. However, the combination of pre-departure screening with test-and-isolate and/or self-quarantine may be sufficient for risk management. Even without any border controls towards multiple G2 and G3 entities (at a constant rate of 1000 imported cases per week), a G2 entity would be able to keep infections under control (albeit at a relatively high level of 14000 new cases per week or 2000 new cases per day). |  |  |

**Table 4.** Quality assessment of modelling studies identified in the updated search.

|  | Questions | Gankin 2021 | Han 2021 | Hossain 2020 | Ip 2021 | Jablonska 2021 | Kwak 2021 |
| --- | --- | --- | --- | --- | --- | --- | --- |
| Model structure | 1. Are the structural assumptions transparent and justified? | Yes | Yes | Yes | Yes | Yes | Yes |
|  | 2. Are the structural assumptions reasonable given the overall objective, perspective, and scope of the model? | Yes | Yes | Yes | Yes | Yes | Yes |
|  | 3. Are the input parameters transparent and justified? | Yes | Yes | Yes | Yes | Yes | Yes |
|  | 4. Are the input parameters reasonable? | Yes | Yes | Yes | Yes | Yes | Yes |
| Validation (external) | 5. Has the external validation process been described? | Yes | Yes | Yes | No | No | Yes |
|  | 6. Has the model been shown to be externally valid? | Yes | Yes | Yes | No | No | Yes |
| Validation (internal) | 7. Has the internal validation process been described? | No | No | No | No | No | No |
|  | 8. Has the model been shown to be internally valid? | No | No | No | No | No | No |
| Uncertainty | 9. Was there an adequate assessment of the effects of uncertainty? | Yes | Yes | No | No | No | No |
| Transparency | 10. Was technical documentation, in sufficient detail to allow (potentially) for replication, made available openly or under agreements that protect intellectual property? | No | No | No | Yes | No | Yes |

|  | Questions | Sun 2021 | Yang 2021 | Yang 2021 | Yu 2021 | Zhu 2021 |
| --- | --- | --- | --- | --- | --- | --- |
| Model structure | 1. Are the structural assumptions transparent and justified? | Yes | Yes | Yes | Yes | Yes |
|  | 2. Are the structural assumptions reasonable given the overall objective, perspective, and scope of the model? | Yes | Yes | Yes | Yes | Yes |
|  | 3. Are the input parameters transparent and justified? | Yes | Yes | Yes | Yes | Yes |
|  | 4. Are the input parameters reasonable? | Yes | Yes | Yes | Yes | Yes |
| Validation (external) | 5. Has the external validation process been described? | No | Yes | Yes | No | No |
|  | 6. Has the model been shown to be externally valid? | No | Yes | Yes | No | No |
| Validation (internal) | 7. Has the internal validation process been described? | No | No | No | No | No |
|  | 8. Has the model been shown to be internally valid? | No | No | No | No | No |
| Uncertainty | 9. Was there an adequate assessment of the effects of uncertainty? | Yes | Yes | Yes | No | No |
| Transparency | 10. Was technical documentation, in sufficient detail to allow (potentially) for replication, made available openly or under agreements that protect intellectual property? | No | No | No | No | No |

For the nonrandomized comparative study,^38^ the individual quality assessment domains are presented in Table 5. Overall, this study received 6 out of 8 stars, making it of good quality.

**Table 5.** Quality assessment of nonrandomized studies identified in the updated search.

|  | **Selection 1** | **Selection 2** | **Selection 3** | **Selection 4** | **Comparability 1** | **Outcome 1** | **Outcome 2** | **Outcome 3** | **Total** |
| --- | --- | --- | --- | --- | --- | --- | --- | --- | --- |
| **Gwee 2021** | 1 | 0 | 1 | 1 | 0 | 1 | 1 | 1 | 6 |

**Table 6.** Quality assessment of modelling studies identified in the original Cochrane review.

|  | Questions | Adekunle 2020 | Anderson 2021 | Anzai 2020 | Banholzer 2021 | Binny 2021 | Boldog 2020 |
| --- | --- | --- | --- | --- | --- | --- | --- |
| Model structure | 1. Are the structural assumptions transparent and justified? | No | Yes | No | Yes | Yes | Yes |
|  | 2. Are the structural assumptions reasonable given the overall objective, perspective, and scope of the model? | No | No | No | No | Yes | Yes |
|  | 3. Are the input parameters transparent and justified? | Yes | Yes | Yes | Yes | Yes | Yes |
|  | 4. Are the input parameters reasonable? | No | No | No | Yes | Yes | No |
| Validation (external) | 5. Has the external validation process been described? | Yes | Yes | Yes | Yes | No | No |
|  | 6. Has the model been shown to be externally valid? | Yes | Yes | Yes | Yes | No | No |
| Validation (internal) | 7. Has the internal validation process been described? | No | No | No | No | No | No |
|  | 8. Has the model been shown to be internally valid? | No | No | No | No | No | No |
| Uncertainty | 9. Was there an adequate assessment of the effects of uncertainty? | Yes | No | No | Yes | Yes | No |
| Transparency | 10. Was technical documentation, in sufficient detail to allow (potentially) for replication, made available openly or under agreements that protect intellectual property? | No | Yes | No | Yes | No | Yes |

|  | Questions | Chen 2020 | Chinazzi 2020 | Costantino 2020 | Davis 2020 | Deeb 2020 | Grannell 2020 |
| --- | --- | --- | --- | --- | --- | --- | --- |
| Model structure | 1. Are the structural assumptions transparent and justified? | Yes | Yes | Yes | Yes | Yes | Yes |
|  | 2. Are the structural assumptions reasonable given the overall objective, perspective, and scope of the model? | Yes | Yes | No | Yes | Yes | No |
|  | 3. Are the input parameters transparent and justified? | Yes | Yes | Yes | No | Yes | No |
|  | 4. Are the input parameters reasonable? | Yes | No | No | No | Yes | Yes |
| Validation (external) | 5. Has the external validation process been described? | Yes | Yes | Yes | Yes | Yes | No |
|  | 6. Has the model been shown to be externally valid? | No | No | No | No | Yes | No |
| Validation (internal) | 7. Has the internal validation process been described? | No | No | No | No | No | No |
|  | 8. Has the model been shown to be internally valid? | No | No | No | No | No | No |
| Uncertainty | 9. Was there an adequate assessment of the effects of uncertainty? | Yes | Yes | Yes | Yes | No | No |
| Transparency | 10. Was technical documentation, in sufficient detail to allow (potentially) for replication, made available openly or under agreements that protect intellectual property? | No | No | No | No | No | No |

|  | Questions | Kang 2020 | Kwok 2021 | Liebig 2021 | Linka 2020a | Linka 2020b | McLure 2021 |
| --- | --- | --- | --- | --- | --- | --- | --- |
| Model structure | 1. Are the structural assumptions transparent and justified? | No | No | No | Yes | Yes | Yes |
|  | 2. Are the structural assumptions reasonable given the overall objective, perspective, and scope of the model? | No | No | No | No | No | Yes |
|  | 3. Are the input parameters transparent and justified? | Yes | No | No | Yes | Yes | Yes |
|  | 4. Are the input parameters reasonable? | No | No | Yes | No | Yes | Yes |
| Validation (external) | 5. Has the external validation process been described? | Yes | No | No | Yes | Yes | No |
|  | 6. Has the model been shown to be externally valid? | Yes | No | No | No | No | No |
| Validation (internal) | 7. Has the internal validation process been described? | No | No | No | No | No | No |
|  | 8. Has the model been shown to be internally valid? | No | No | No | No | No | No |
| Uncertainty | 9. Was there an adequate assessment of the effects of uncertainty? | No | No | No | No | No | No |
| Transparency | 10. Was technical documentation, in sufficient detail to allow (potentially) for replication, made available openly or under agreements that protect intellectual property? | No | No | No | No | No | Yes |

|  | Questions | Nakamura<br>2020 | Nowrasteh<br>2020 | Odendaal<br>2020 | Pinotti 2020 | Russell 2021 | Shi 2020 |
| --- | --- | --- | --- | --- | --- | --- | --- |
| Model structure | 1. Are the structural assumptions transparent and justified? | No | No | No | Yes | Yes | Yes |
|  | 2. Are the structural assumptions reasonable given the overall objective, perspective, and scope of the model? | No | No | No | Yes | Yes | No |
|  | 3. Are the input parameters transparent and justified? | Yes | Yes | Yes | Yes | Yes | Yes |
|  | 4. Are the input parameters reasonable? | No | No | Yes | Yes | Yes | Yes |
| Validation<br>(external) | 5. Has the external validation process been described? | No | Yes | Yes | Yes | Yes | Yes |
|  | 6. Has the model been shown to be externally valid? | No | Yes | No | Yes | No | No |
| Validation<br>(internal) | 7. Has the internal validation process been described? | No | No | No | No | No | No |
|  | 8. Has the model been shown to be internally valid? | No | No | No | No | No | No |
| Uncertainty | 9. Was there an adequate assessment of the effects of uncertainty? | No | Yes | No | No | No | No |
| Transparency | 10. Was technical documentation, in sufficient detail to allow (potentially) for replication, made available openly or under agreements that protect intellectual property? | No | Yes | No | No | Yes | Yes |

|  | Questions | Sruthi 2020 | Utsunomiya 2020 | Wells 2020 | Yang 2020 | Zhang 2021 | Zhang 2020 |
| --- | --- | --- | --- | --- | --- | --- | --- |
| Model structure | 1. Are the structural assumptions transparent and justified? | No | Yes | Yes | Yes | Yes | No |
|  | 2. Are the structural assumptions reasonable given the overall objective, perspective, and scope of the model? | No | No | Yes | Yes | No | No |
|  | 3. Are the input parameters transparent and justified? | Yes | Yes | Yes | Yes | Yes | Yes |
|  | 4. Are the input parameters reasonable? | Yes | Yes | No | Yes | Yes | Yes |
| Validation (external) | 5. Has the external validation process been described? | Yes | Yes | Yes | Yes | No | Yes |
|  | 6. Has the model been shown to be externally valid? | No | Yes | Yes | No | No | No |
| Validation (internal) | 7. Has the internal validation process been described? | No | Yes | No | No | Yes | No |
|  | 8. Has the model been shown to be internally valid? | No | Yes | No | No | Yes | No |
| Uncertainty | 9. Was there an adequate assessment of the effects of uncertainty? | No | No | Yes | Yes | No | No |
| Transparency | 10. Was technical documentation, in sufficient detail to allow (potentially) for replication, made available openly or under agreements that protect intellectual property? | Yes | Yes | Yes | Yes | Yes | No |

|  | Questions | Zhong 2021 |
| --- | --- | --- |
| Model structure | 1. Are the structural assumptions transparent and justified? | No |
|  | 2. Are the structural assumptions reasonable given the overall objective, perspective, and scope of the model? | Yes |
|  | 3. Are the input parameters transparent and justified? | Yes |
|  | 4. Are the input parameters reasonable? | No |
| Validation (external) | 5. Has the external validation process been described? | Yes |
|  | 6. Has the model been shown to be externally valid? | No |
| Validation (internal) | 7. Has the internal validation process been described? | Yes |
|  | 8. Has the model been shown to be internally valid? | Yes |
| Uncertainty | 9. Was there an adequate assessment of the effects of uncertainty? | No |
| Transparency | 10. Was technical documentation, in sufficient detail to allow (potentially) for replication, made available openly or under agreements that protect intellectual property? | No |

### Travel restrictions reducing or stopping cross-border travel

Similar to the Cochrane review,^3^ we did not attempt to differentiate between a complete border closure versus travel restrictions (leading to varying degrees of difficulty in crossing borders), and instead report these in a combined intervention category. Studies reported on (1) ‘cases avoided due to the measure’ (n = 30),^6–10, 12–14, 16, 18–21, 23–25, 28, 29, 32–39, 41, 42, 46, 47^ (2) the shift in epidemic development (n = 19),^8, 10, 11, 14, 15, 17, 18, 21, 22, 26, 27, 30, 31, 36, 40, 42, 44, 45, 48^ (3) cases detected due to the measure (n = 3),^43, 46, 48^ and (4) secondary outcomes (n = 1).^19^ The newly identified studies add to the body of the evidence, but do not change the conclusions of the Cochrane review;^3^ usually positive direction of effect with travel restrictions/ border closures (low to very low-certainty evidence).

#### 1. Cases avoided due to the measure

This outcome is subclassified into:

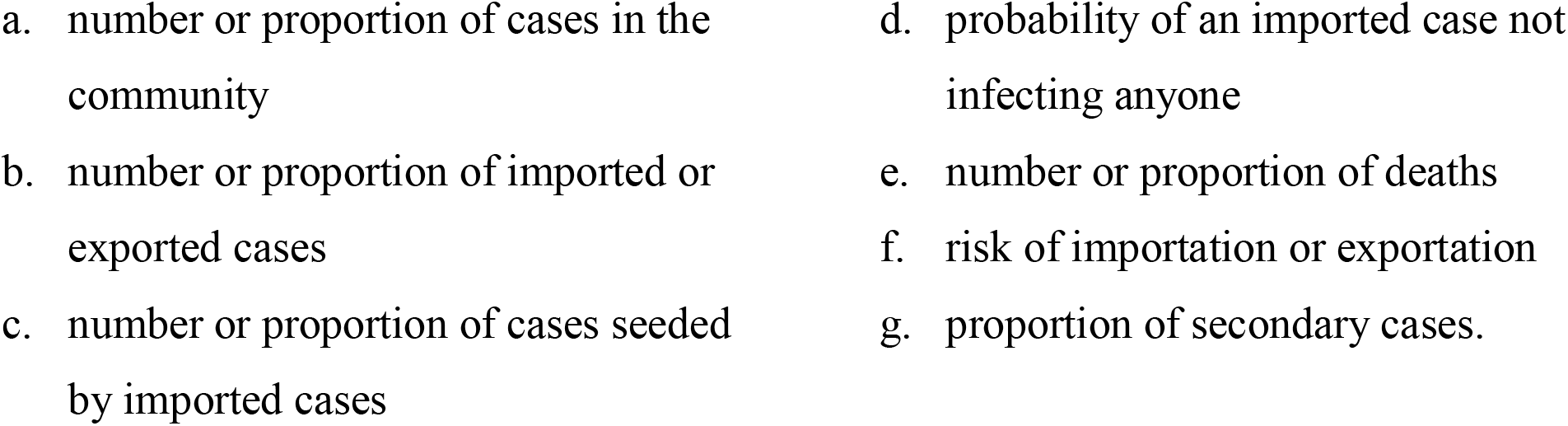

|  |  |
| --- | --- |
| a. number or proportion of cases in the community | d. probability of an imported case not infecting anyone |
| b. number or proportion of imported or exported cases | e. number or proportion of deaths |
| c. number or proportion of cases seeded by imported cases | f. risk of importation or exportation |
|  | g. proportion of secondary cases. |

a. Number or proportion of cases in the community Five new studies^37, 38, 41, 42, 47^ was identified in the updated search. In total, we included 17 modelling studies,^7, 9, 10, 12, 14, 16, 18, 19, 21, 25, 33, 35–38, 41, 42, 47^ with most (n = 13) reporting fewer new cases (1.8%^10^ to 97.8%^18^) with travel restrictions; four studies^25, 33, 36, 37^ reported no significant positive effect (very low certainty evidence).
b. Number or proportion of imported or exported cases Three new studies^38, 39, 46^ were identified in the updated search. In total, we included 12 modelling studies,^6, 8, 12–14, 20, 23, 28, 32, 38, 39, 46^ with most (n = 11) reporting decreased cases (18%^20^ to 99%^12^ reductions) with travel restrictions; one study^6^ reported mixed results (very low certainty evidence).
c. Number or proportion of cases seeded by imported cases No studies were identified by the Cochrane review^3^ or in the updated search.
d. Probability of an imported case not infecting anyone No studies were identified by the Cochrane review^3^ or in the updated search.
e. Number or proportion of deaths <u>No new studies were identified in the updated search</u>. The Cochrane review^3^ identified three modelling studies^10, 14, 19^ with all reporting decreased deaths (4.3%^10^ to 98%^14^) with travel restrictions (very low certainty evidence).
f. Risk of importation or exportation <u>No new studies were identified in the updated search.</u> The Cochrane review^3^ identified three modelling studies.^24, 29, 34^ Two studies^24, 34^ reported decreased risk with travel restrictions; however, no direct effect estimates were reported. One study^29^ reported mixed results (very low certainty evidence); lessening restrictions led to an increased risk of importation at some airports, but a decreased risk at other airports.
g. Proportion of secondary cases No studies were identified by the Cochrane review^3^ or in the updated search.

#### 2. Shift in epidemic development

This outcome is subclassified into:

|  |  |
| --- | --- |
| a. probability of eliminating the epidemic | e. number or proportion of cases at peak |
| b. effective reproduction number | f. median time to peak |
| c. time to outbreak | g. epidemic curve peak |
| d. risk of an outbreak | h. epidemic growth acceleration |
|  | i. exportation growth rate. |

a. Probability of eliminating the epidemic <u>No new studies were identified in the updated search</u>. The Cochrane review^3^ identified one modelling study^10^ reporting mixed results (very low-certainty evidence).
b. Effective reproduction number (Rt) One new study^40^ was identified in the updated search. In total, we included three modelling studies,^21, 30, 40^ with most (n = 2) reporting benefit with travel restrictions; one study^30^ reported mixed results (dependent on the severity of the border closures) (very low-certainty evidence). The newest-identified study^40^ reported a beneficial change under a low reproduction number (Ro) of 1.4. The border control measure would have reduced 90% of the passenger numbers and gained an extra 32.5 days of outbreak arrival time. The effect of border control was weaker under a medium Ro (1.68); nevertheless, it still resulted in 20.0 extra days under the same control level. However, under a high Ro (2.92), only 10 extra days were obtained.
c. Time to outbreak Four new studies^18, 44, 45, 48^ were identified in the updated search. In total, we included 10 modelling studies,^8, 15, 17, 18, 22, 26, 36, 44, 45, 48^ with most reporting benefits (e.g., <1 day^8^ to 26 weeks^48^) with travel restrictions; three studies^17, 36, 44^ reported mixed results (very low-certainty evidence).
d. Risk of an outbreak <u>No new studies were identified in the updated search</u>. The Cochrane review^3^ identified two modelling studies^8,11^ with one study^8^ reporting benefits (1% to 37%) with travel restrictions, while the second study^11^ reported mixed results (very low-certainty evidence).
e. Number or proportion of cases at peak <u>No new studies were identified in the updated search</u>. The Cochrane review^3^ identified two modelling studies;^10, 17^ one study^17^ reported benefits with travel restrictions (0.3% to 8% at peak) and the other study^10^ reported that an early implementation of border restrictions or a delayed border closure would lead to 79 (95% CI 67 to 97) and 91 (95% CI 77 to 110) daily cases at the epidemic peak, respectively (low-certainty evidence).
f. Median time to peak One new study^42^ was identified in the updated search. This modelling study^42^ assessed the time to peak which equalled 31 days from the first death reported in each country, with a comparable mean (31.32 ± 13.94 days) (low-certainty evidence). In the multivariate analysis, border closure day was significantly associated with time to peak deaths (0.297).
g. Epidemic curve peak One new study^14^ was identified in the updated search. This modelling study^14^ assessed that travel restrictions will delay the epidemic curve about 50 days in time (low-certainty evidence).
h. Epidemic growth acceleration <u>No new studies were identified in the updated search</u>. The Cochrane review^3^ identified one modelling study^31^ reporting benefits from travel restrictions (−6.05% change) (low-certainty evidence).
i. Exportation growth rate <u>No new studies were identified in the updated search</u>. The Cochrane review^3^ identified one modelling study^27^ reporting benefits from travel restrictions (within China) (low-certainty evidence).

#### 3. Cases detected due to the measure

This outcome is subclassified into:

|  |  |
| --- | --- |
| a. days at risk of transmission | c. positive predictive value (PPV) |
| b. number or proportion of cases detected | d. probability of releasing an infected individual into the community. |

a. Days at risk of transmission No studies were identified by the Cochrane review^3^ or in the updated search.
b. Number or proportion of cases detected Two new modelling studies^43, 48^ were identified in the updated search. These studies reported mixed effects in terms of the proportion of cases detected. The first study^43^ investigated the effect of international travel restrictions by difference in policy levels between the government and agent in 216 countries. It was found that earlier agent lockdowns (when compared to government lockdown policy) were associated with a more rapid acceleration in cases. In contrast, earlier government closure of the border compared to the agent was not necessarily associated with a slower acceleration of cases. These results indicated that local transmission may have a greater effect on disease burden compared to imported cases. The second study^48^ assessed the effect of border control policies, in combination with internal measures (model with built-in imported risk and [1-tier] contact tracing). The curve remained flattened in G2 countries, provided that there was a sufficient confining domestic control policy (R<1 without the imported cases) and that the imported risk was not increasing over time. In a scenario in which two G2 entities had completely open borders toward each other and no quarantine or screening policies, both entities experienced decreasing numbers of cases, similarly to the use of a closed border policy. Both entities were also able to withstand the risk of importation and maintain a decreasing trend for the number of newly identified cases and hospitalized patients over time if the number of infected people upon arrival was doubled. This conclusion could not be made for a G2 entity that opened its border to a G3 entity. The effort of the G2 entity to contain the COVID-19 outbreak was jeopardized by increasing the number of imported cases from G3. Additional border control policies would be required for travelers from G3 entities. Therefore, individual measures when implemented alone may not be able to avoid another lock down within half a year if vaccines are still not widely deployed. However, the combination of pre-departure screening with test-and-isolate and/or self-quarantine may be sufficient for risk management. Even without any border controls towards multiple G2 and G3 entities (at a constant rate of 1000 imported cases per week), a G2 entity would be able to keep infections under control (albeit at a relatively high level of 14000 new cases per week or 2000 new cases per day).
c. Positive predictive value (PPV) No studies were identified by the Cochrane review^3^ or in the updated search.
d. Probability of releasing an infected individual into the community <u>One study was identified in the updated search</u>.^46^ This study investigated the effects of a travel ban for non-Hong Kong residents from overseas and testing and quarantine for those permitted to travel. The results indicated that tightening travel measures to 21-day quarantine reduced the risk of releasing infectious travellers to 0 for all examined countries or regions.

#### 4. Secondary outcomes

This outcome is subclassified into:

|  |  |
| --- | --- |
| a. infectious disease transmission outcomes | d. adverse effects (if identified in studies assessing at least one primary outcome). |
| b. healthcare utilisation |  |
| c. resource requirements |  |

a. Infectious disease transmission outcomes No studies were identified by the Cochrane review^3^ or in the updated search.
b. Healthcare utilisation <u>No new studies were identified in the updated search.</u> The Cochrane review^3^ identified one modelling study^19^ reporting benefits from travel restrictions on secondary outcomes related to healthcare utilisation. They showed that border closures alone, while beneficial, can not prevent hospitals from eventually reaching their capacity.
c. Resource requirements No studies were identified by the Cochrane review^3^ or in the updated search.
d. Adverse effects (if identified in studies assessing at least one primary outcome) No studies were identified by the Cochrane review^3^ or in the updated search.

## Conclusions

In addition to the 31 studies identified by the Cochrane review, we identified 12 additional studies, mostly modelling studies (e.g., simulated border closures), that compared the effectiveness of limiting travel as a measure to reduce the spread of COVID-19. The added studies did not change the main conclusions of the Cochrane review nor the quality of the evidence (very low to low certainty). However, it did add to the evidence base for most outcomes and provided evidence for ‘reproduction number’ and ‘number or proportion of cases detected’ that were not available in the Cochrane review^3^. Due to the nature of GRADEing the certainty of the evidence, it is unlikely that more studies will change the quality of the evidence.

In conclusion, weak evidence supports the use of border closures to limit the spread of COVID-19 from region to regions. Real-world studies are required to support these conclusions.

## Data Availability

All data produced in the present work are contained in the manuscript.

## Appendix 1. Details for quality assessment decisions

### 1. Gankin, Infect Genet Evol, 2021

|  | Questions | Decision | Reason |
| --- | --- | --- | --- |
| Model structure | 1. Are the structural assumptions transparent and justified? | Yes | No to minor concerns. Stochastic model. |
|  | 2. Are the structural assumptions reasonable given the overall objective, perspective and scope of the model? | Yes | No to minor concerns. |
|  | 3. Are the input parameters transparent and justified? | Yes | No to minor concerns. Input parameters were justified. |
|  | 4. Are the input parameters reasonable? | Yes | No to minor concerns. Input parameters were probably reasonable. |
| Validation (external) | 5. Has the external validation process been described? | Yes | No to minor concerns. |
|  | 6. Has the model been shown to be externally valid? | Yes | No to minor concerns. The reported and model-produced data from April 22, 2020, to July 12, 2020 were used for model calibration, and from July 13, 2020 to August 1, 2020 – for model validation. The data before April 22, 2020, were used solely for the initial conditions to increase the model fit robustness, since the initial number of cases was relatively small in comparison to subsequent periods. The August 1, 2020, has been selected as the end date of our simulations to agree with the dates of genomic analysis based on available analyzed SARS-CoV-2 sequences collection times. |
| Validation (internal) | 7. Has the internal validation process been described? | No | Not reported. |
|  | 8. Has the model been shown to be internally valid? | No | Moderate concerns |
| Uncertainty | 9. Was there an adequate assessment of the effects of uncertainty? | Yes | No to minor concerns. |
| Transparency | 10. Was technical documentation, in sufficient detail to allow (potentially) for replication, made available openly or under agreements that protect intellectual property? | No | Moderate concerns. The uncertainty estimates for the stochastic estimates are wider, which may be since stochastic model has more parameters and higher variability in the outputs while phylodynamic models has strong priors. Even so, the code use to run the model was not provided. |

### 2. Han, Proc Natl Acad Sci USA, 2021

|  | Questions | Decision | Reason |
| --- | --- | --- | --- |
| Model structure | 1. Are the structural assumptions transparent and justified? | Yes | No to minor concerns. Different change points for each of the transmissibility parameters were allowed for and identified via a Bayesian approach. |
|  | 2. Are the structural assumptions reasonable given the overall objective, perspective and scope of the model? | Yes | No to minor concerns. See above. |
|  | 3. Are the input parameters transparent and justified? | Yes | No to minor concerns. The number of daily newly confirmed cases of origin k on day t were sourced from sourced from the European Centre for Disease Prevention and Control. |
|  | 4. Are the input parameters reasonable? | Yes | No to minor concerns. See above. |
| Validation (external) | 5. Has the external validation process been described? | Yes | To validate the linear model, the authors ran an estimation assuming a conditional Poisson distribution on $y_{it}$'s, using a sub-sample between April 8th to April 28th, the period during which the total newly confirmed cases for all cities become non-negative. |
|  | 6. Has the model been shown to be externally valid? | Yes | No to minor concerns. |
| Validation (internal) | 7. Has the internal validation process been described? | No | Not reported. |
|  | 8. Has the model been shown to be internally valid? | No | Moderate concerns. |
| Uncertainty | 9. Was there an adequate assessment of the effects of uncertainty? | Yes | No to minor concerns. To alleviate the concern about the uncertainty of incubation period and reporting delay, the authors conducted some robustness checks using a 7-day lag centered average and a 14-day lag centered average, respectively. |
| Transparency | 10. Was technical documentation, in sufficient detail to allow (potentially) for replication, made available openly or under agreements that protect intellectual property? | No | Moderate concerns. The code use to run the model was not provided. |

### 3. Hossain, Epidemics, 2020

|  | Questions | Decision | Reason |
| --- | --- | --- | --- |
| Model structure | 1. Are the structural assumptions transparent and justified? | Yes | No to minor concerns. Based on the Susceptible, Infected, and Recovered (SIR) meta-population model. |
|  | 2. Are the structural assumptions reasonable given the overall objective, perspective and scope of the model? | Yes | No to minor concerns. "...assumed that infected cases could pass the border screening or move to another location only during their incubation period (referred to as exposed cases)." |
|  | 3. Are the input parameters transparent and justified? | Yes | No to minor concerns. "Airline passenger data were collected from the International Air Transport Association database." |
|  | 4. Are the input parameters reasonable? | Yes | No to minor concerns. "We collected the actual passenger data for the top 10 visiting cities leaving from Wuhan Tianhe International Airport before the lockdown of Wuhan city from 30 Dec to 20 Jan, 2020. Note that we did not have data for railroad and other forms of transport, and thus made an assumption that the total number of travelers is 4 times higher than that of air transport except for certain cities on Hainan Island that have no road connections to Wuhan. We made this assumption because that the number of train passengers is few times higher than that of airline in China (Statista, 2018) and the results from a population migration database suggested a similar ratio (The Tencent database, 2020)." |
| Validation (external) | 5. Has the external validation process been described? | Yes | No to minor concerns. |
|  | 6. Has the model been shown to be externally valid? | Yes | The predicted reporting delay was very close to the actual reporting lag. Overall, the predicted cumulative number of both imported and secondary cases after adjusted by the reporting lag time of top 10 visiting cities demonstrated a similar increasing trend in cumulative numbers of confirmed cases during each early emergence period. |
| Validation (internal) | 7. Has the internal validation process been described? | No | Not reported. |
|  | 8. Has the model been shown to be internally valid? | No | Moderate concerns. |
| Uncertainty | 9. Was there an adequate assessment of the effects of uncertainty? | No | Major concerns. Not reported. |
| Transparency | 10. Was technical documentation, in sufficient detail to allow (potentially) for replication, made available openly or under agreements that protect intellectual property? | No | Moderate concerns. The code use to run the model was not provided. |

### 4. Ip, Int J Environ Res Public Health, 2021

|  | Questions | Decision | Reason |
| --- | --- | --- | --- |
| Model structure | 1. Are the structural assumptions transparent and justified? | Yes | No to minor concerns. GSTARX was based on previous studies. |
|  | 2. Are the structural assumptions reasonable given the overall objective, perspective and scope of the model? | Yes | No to minor concerns. See above. |
|  | 3. Are the input parameters transparent and justified? | Yes | No to minor concerns. COVID-19 case data from 25 January 2020 to 12 September 2020 were obtained from the webpage of the Department of Health, Australia Government and the Australian Bureau of Statistics. |
|  | 4. Are the input parameters reasonable? | Yes | No to minor concerns. See above. |
| Validation (external) | 5. Has the external validation process been described? | No | Not reported. |
|  | 6. Has the model been shown to be externally valid? | No | Moderate concerns. |
| Validation (internal) | 7. Has the internal validation process been described? | No | Not reported. |
|  | 8. Has the model been shown to be internally valid? | No | Moderate concerns. |
| Uncertainty | 9. Was there an adequate assessment of the effects of uncertainty? | No | Major concerns. Not reported. |
| Transparency | 10. Was technical documentation, in sufficient detail to allow (potentially) for replication, made available openly or under agreements that protect intellectual property? | Yes | No to minor concerns. |

### 5. Jablonska, medRxiv, 2021

|  | Questions | Decision | Reason |
| --- | --- | --- | --- |
| Model structure | 1. Are the structural assumptions transparent and justified? | Yes | No to minor concerns. Loess regression based on prior literature. |
|  | 2. Are the structural assumptions reasonable given the overall objective, perspective and scope of the model? | Yes | No to minor concerns. See above. |
|  | 3. Are the input parameters transparent and justified? | Yes | No to minor concerns. "Data on COVID-19 deaths, infections, the number of tests, bed capacity, government restrictions, population size and urban population size were taken from the Institute for Health Metrics and Evaluation. Missing dates of government restrictions, if officially issued, were found on Wikipedia. Mobility scores were uploaded from Google COVID-19 Community Mobility Reports" |
|  | 4. Are the input parameters reasonable? | Yes | No to minor concerns. See above. |
| Validation (external) | 5. Has the external validation process been described? | No | Not reported. |
|  | 6. Has the model been shown to be externally valid? | No | Moderate concerns. |
| Validation (internal) | 7. Has the internal validation process been described? | No | Not reported. |
|  | 8. Has the model been shown to be internally valid? | No | Moderate concerns. |
| Uncertainty | 9. Was there an adequate assessment of the effects of uncertainty? | No | No to minor concerns. Sensitivity analyses were performed using a variety of selection algorithms: stepwise, backward, forward and genetic algorithm, to limit the number of covariates, increase model precision and improve model fit. |
| Transparency | 10. Was technical documentation, in sufficient detail to allow (potentially) for replication, made available openly or under agreements that protect intellectual property? | No | Moderate concerns. The code use to run the model was not provided. |

### 6. Kwak, PloS one, 2021

|  | Questions | Decision | Reason |
| --- | --- | --- | --- |
| Model structure | 1. Are the structural assumptions transparent and justified? | Yes | No to minor concerns. Use of reinforcement learning agent and seeking of an optimal policy with the Dueling Double Deep Q-Network and use of SIRD model was based on prior literature. |
|  | 2. Are the structural assumptions reasonable given the overall objective, perspective and scope of the model? | Yes | No to minor concerns. See above. |
|  | 3. Are the input parameters transparent and justified? | Yes | No to minor concerns. Use of real-world data: "For each country and territory, index case date (date of the first locally confirmed patient), the numbers tested, confirmed infection, recovered and dead were collected from Johns Hopkins coronavirus data repository, Centers for Disease Control and Prevention's reports and WHO's case reports. Population size, population density, population mid-year (aged 15 to 65 years old), gross domestic product (GDP), geological information (longitude, latitude) and life expectancy from the United Nations database, Wikipedia, and official announcements through the news were used in our algorithm for the country and territory specific population characteristics and healthcare setting" |
|  | 4. Are the input parameters reasonable? | Yes | No to minor concerns. See above. |
| Validation (external) | 5. Has the external validation process been described? | Yes | No to minor concerns. |
|  | 6. Has the model been shown to be externally valid? | Yes | No to minor concerns. "For policy validation, we used an evaluation technique to estimate how the differences between the government and agent policies relate to accelerated infection, death and recovery cases [32, 33]. The total acceleration was calculated and derived in relation to the difference at the policy level. In general, earlier agent lockdowns when compared to government lockdown policy was related to a more rapid acceleration in cases" |
| Validation (internal) | 7. Has the internal validation process been described? | No | Not reported. |
|  | 8. Has the model been shown to be internally valid? | No | Moderate concerns. |
| Uncertainty | 9. Was there an adequate assessment of the effects of uncertainty? | No | Major concerns. Not reported. |
| Transparency | 10. Was technical documentation, in sufficient detail to allow (potentially) for replication, made available openly or under agreements that protect intellectual property? | Yes | No to minor concerns. |

### 7. Sun, Transp Res Part A Policy Pract, 2021

|  | Questions | Decision | Reason |
| --- | --- | --- | --- |
| Model structure | 1. Are the structural assumptions transparent and justified? | Yes | No to minor concerns. Susceptible–Exposed–Infectious–Recovered (SEIR) model extended onto a network structure, based on the concepts of the GLobal Epidemic and Mobility (GLEaM) Model |
|  | 2. Are the structural assumptions reasonable given the overall objective, perspective and scope of the model? | Yes | No to minor concerns. See above. |
|  | 3. Are the input parameters transparent and justified? | Yes | No to minor concerns. Based on reported cases of variants from https://cov-lineages.org. Real word commuting flow data obtained from prior study. |
|  | 4. Are the input parameters reasonable? | Yes | No to minor concerns. See above. |
| Validation (external) | 5. Has the external validation process been described? | No | Not reported. |
|  | 6. Has the model been shown to be externally valid? | No | Moderate concerns. |
| Validation (internal) | 7. Has the internal validation process been described? | No | Not reported. |
|  | 8. Has the model been shown to be internally valid? | No | Moderate concerns. |
| Uncertainty | 9. Was there an adequate assessment of the effects of uncertainty? | Yes | No to minor concerns. Sensitivity analysis on variable, ranging from 0.00 (equal to not cutting any flights) and 0.99 (equal to cutting all flights by 99%). |
| Transparency | 10. Was technical documentation, in sufficient detail to allow (potentially) for replication, made available openly or under agreements that protect intellectual property? | No | Moderate concerns. The code use to run the model was not provided. |

### 8. Yang, Emerg Infect Dis, 2021

|  | Questions | Decision | Reason |
| --- | --- | --- | --- |
| Model structure | 1. Are the structural assumptions transparent and justified? | Yes | No to minor concerns. Use of previously established Kaplan–Meier survival curve and accelerated failure time model. |
|  | 2. Are the structural assumptions reasonable given the overall objective, perspective and scope of the model? | Yes | No to minor concerns. See above. |
|  | 3. Are the input parameters transparent and justified? | Yes | No to minor concerns. Publicly available case data from Jan 1 to July 31, 2020 were used. |
|  | 4. Are the input parameters reasonable? | Yes | No to minor concerns. See above. |
| Validation (external) | 5. Has the external validation process been described? | Yes | "For some countries had many fluctuations over the study period (e.g., Guyana) or experienced a much larger outbreak during the later of our study period (e.g., Argentina), which could result in right-censoring for the first epidemic peak (i.e., no epidemic peak was observed according to the peak measurement). To validate if our results would be greatly affected by these issues, we used the reaching a certain threshold for cumulative incidence. We believe using the alternative outcome measurements could overcome the abovementioned misclassifications or right-censoring issues, although for some countries with low COVID-19 circulation it could introduce right-censoring as well" |
|  | 6. Has the model been shown to be externally valid? | Yes | No to minor concerns. |
| Validation (internal) | 7. Has the internal validation process been described? | No | Not reported. |
|  | 8. Has the model been shown to be internally valid? | No | Moderate concerns. |
| Uncertainty | 9. Was there an adequate assessment of the effects of uncertainty? | Yes | No to minor concerns. Authors "performed a sensitivity analysis by fitting the AFT and Cox models with data that excluding Asian countries, where tended to have stricter enactment, higher adherence and more precautious when implementing these control measures" |
| Transparency | 10. Was technical documentation, in sufficient detail to allow (potentially) for replication, made available openly or under agreements that protect intellectual property? | No | Moderate concerns. The code use to run the model was not provided. |

### 9. Yang, Lancet Reg Health West Pac, 2021

|  | Questions | Decision | Reason |
| --- | --- | --- | --- |
| Model structure | 1. Are the structural assumptions transparent and justified? | Yes | No to minor concerns. A Bayesian framework was used to model the disease history of infected travellers and the travel measures that were in force. |
|  | 2. Are the structural assumptions reasonable given the overall objective, perspective and scope of the model? | Yes | No to minor concerns. Use of previously established model. |
|  | 3. Are the input parameters transparent and justified? | Yes | No to minor concerns. Data on laboratory-confirmed COVID-19 cases whose exposure risk was listed as travel outside Hong Kong were obtained from the Department of Health. Assumption that 30% of symptomatic cases who developed symptoms before travel would board (counter to travel requirements) was based on previous studies. Assumption that test sensitivity for asymptomatic individuals was 62% that of symptomatic individuals was based on a prior study. |
|  | 4. Are the input parameters reasonable? | Yes | No to minor concerns. See above. |
| Validation (external) | 5. Has the external validation process been described? | Yes | To validate the model, the authors conducted a simulation study to illustrate that the estimation algorithm could provide unbiased estimates of parameters. The model recovered the true values for travel volume and prevalence in 96% and 94% of simulations, respectively. |
|  | 6. Has the model been shown to be externally valid? | Yes | No to minor concerns. |
| Validation (internal) | 7. Has the internal validation process been described? | No | Not reported. |
|  | 8. Has the model been shown to be internally valid? | No | Moderate concerns. |
| Uncertainty | 9. Was there an adequate assessment of the effects of uncertainty? | Yes | No to minor concerns. For the sensitivity analysis, the authors used the time-varying test sensitivity from an alternative source to repeat the estimations of COVID-19 prevalence among inbound travellers and the importation risks across different control regimens. |
| Transparency | 10. Was technical documentation, in sufficient detail to allow (potentially) for replication, made available openly or under agreements that protect intellectual property? | No | Moderate concerns. The code use to run the model was not provided. |

### 10. Yu, Int J Environ Res Public Health, 2021

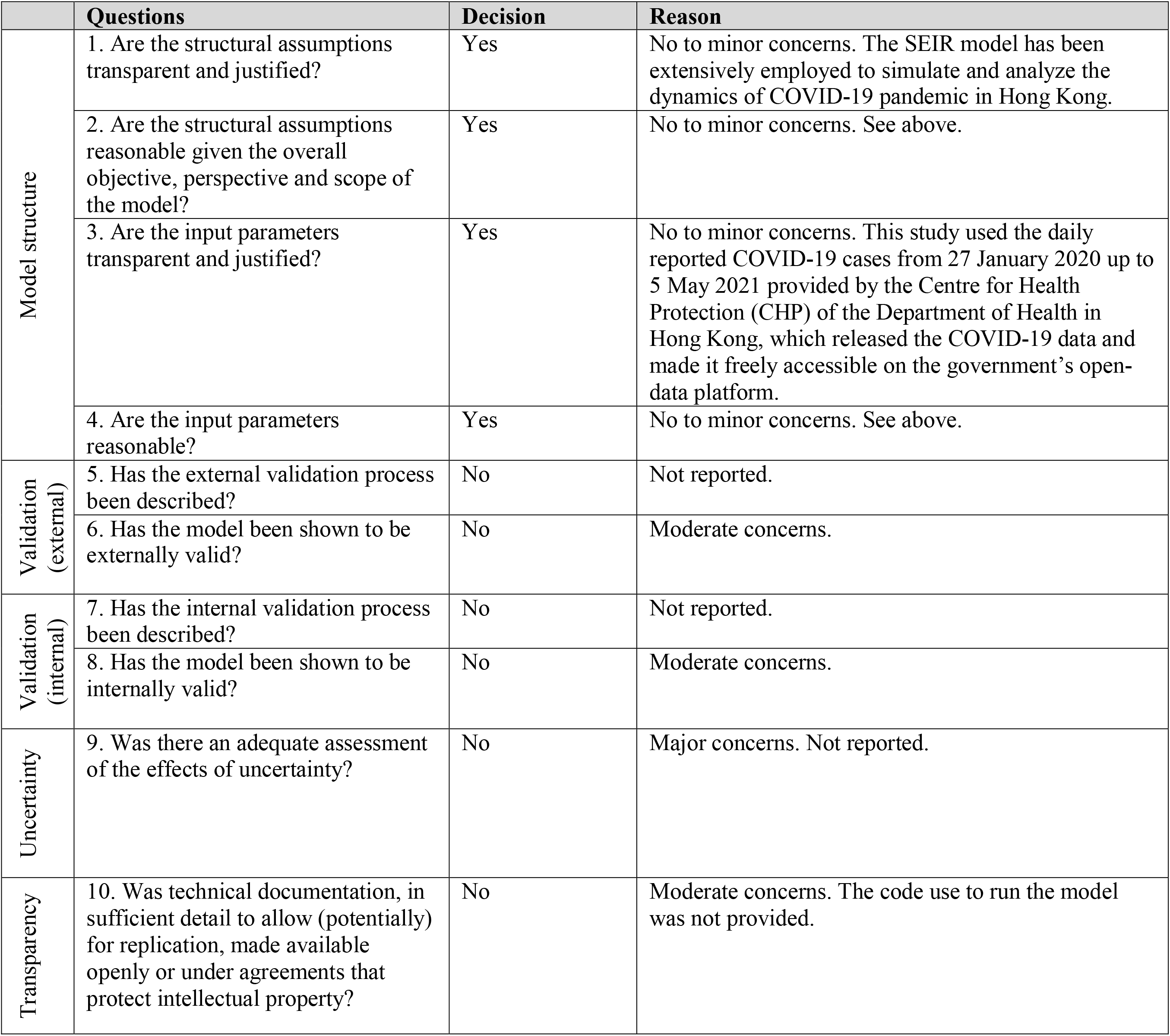

|  | Questions | Decision | Reason |
| --- | --- | --- | --- |
| Model structure | 1. Are the structural assumptions transparent and justified? | Yes | No to minor concerns. The SEIR model has been extensively employed to simulate and analyze the dynamics of COVID-19 pandemic in Hong Kong. |
|  | 2. Are the structural assumptions reasonable given the overall objective, perspective and scope of the model? | Yes | No to minor concerns. See above. |
|  | 3. Are the input parameters transparent and justified? | Yes | No to minor concerns. This study used the daily reported COVID-19 cases from 27 January 2020 up to 5 May 2021 provided by the Centre for Health Protection (CHP) of the Department of Health in Hong Kong, which released the COVID-19 data and made it freely accessible on the government's open-data platform. |
|  | 4. Are the input parameters reasonable? | Yes | No to minor concerns. See above. |
| Validation (external) | 5. Has the external validation process been described? | No | Not reported. |
|  | 6. Has the model been shown to be externally valid? | No | Moderate concerns. |
| Validation (internal) | 7. Has the internal validation process been described? | No | Not reported. |
|  | 8. Has the model been shown to be internally valid? | No | Moderate concerns. |
| Uncertainty | 9. Was there an adequate assessment of the effects of uncertainty? | No | Major concerns. Not reported. |
| Transparency | 10. Was technical documentation, in sufficient detail to allow (potentially) for replication, made available openly or under agreements that protect intellectual property? | No | Moderate concerns. The code use to run the model was not provided. |

### 11. Zhu, Travel medicine and infectious disease, 2021

|  | Questions | Decision | Reason |
| --- | --- | --- | --- |
| Model structure | 1. Are the structural assumptions transparent and justified? | Yes | No to minor concerns. "Extension to the Markov transition based SIR and SEIR models that have long been used to model epidemic spreading and control measures such as lockdowns and vaccines" |
| | 2. Are the structural assumptions reasonable given the overall objective, perspective and scope of the model? | Yes | No to minor concerns. "In general, unidentified and identified infectious, and recovered individuals can either be free or isolated/quarantined. However, with our focus on reopening phases, we optimistically assume that anyone who has been identified as infectious will be isolated immediately to avoid virus spreading. Hospitalized people are also automatically isolated in the model. Assuming only people in the state of unidentified infectious and free to move can travel, only a small percentage of travelers are infectious upon arrival. This percentage, denoted by $a$, is mainly determined by the COVID-19 severity at the origin and the pre-travel screening policies (e.g., people must provide RNA/antibody test results prior to travel)." |
| | 3. Are the input parameters transparent and justified? | Yes | No to minor concerns. "We use $T = 0.91$ with mandatory test for everyone upon arrival [25]. In the absence of such a test, $T = 0$." |
|  | 4. Are the input parameters reasonable? | Yes | No to minor concerns. Table 1- Transitional probability matrix estimation values based on prior literature. |
| Validation (external) | 5. Has the external validation process been described? | No | Not reported. |
|  | 6. Has the model been shown to be externally valid? | No | Moderate concerns. |
| Validation (internal) | 7. Has the internal validation process been described? | No | Not reported. |
|  | 8. Has the model been shown to be internally valid? | No | Moderate concerns. |
| Uncertainty | 9. Was there an adequate assessment of the effects of uncertainty? | No | Major concerns. Not reported. |
| Transparency | 10. Was technical documentation, in sufficient detail to allow (potentially) for replication, made available openly or under agreements that protect intellectual property? | No | Moderate concerns. The code use to run the model was not provided. |

